# Triglyceride Glucose Index Predicts Risk of Adverse Cardio-metabolic and Mortality Outcomes Among Chinese Adults: A Territory-Wide Longitudinal Study

**DOI:** 10.1101/2021.01.04.21249208

**Authors:** Jiandong Zhou, Sharen Lee, Jeremy Hui, Wing Tak Wong, Keith SK Leung, Abraham KC Wai, Tong Liu, Bernard Man Yung Cheung, Gary Tse, Qingpeng Zhang

**Author notes:** Correspondence to: *Prof. Gary Tse PhD FRCP*, Tianjin Key Laboratory of Ionic-Molecular Function of Cardiovascular Disease, Department of Cardiology, Tianjin Institute of Cardiology, Second Hospital of Tianjin Medical University, Tianjin 300211, China, Faculty of Health and Medical Sciences, University of Surrey, GU2 7AL, Guildford, United Kingdom, *Prof. Qingpeng Zhang PhD*, School of Data Science, City University of Hong Kong, Hong Kong, China.

## Abstract

**Background:** The triglyceride glucose (TyG) index has been proposed to be a surrogate of insulin resistance. In the study, we aimed to examine the relationship between TyG index and the risk of new onset DM and secondary outcomes included atrial fibrillation (AF), heart failure (HF), acute myocardial infarction (AMI), ventricular tachycardia/fibrillation (VTF), cardiovascular mortality (CAD) and all-cause mortality.

**Methods:** The retrospective observational study analyzed patients recruited from 1^st^ January 2000 to 31^st^ December 2003 and followed up until 31^st^ December 2019. Demographics, past comorbidities, medications and laboratory tests were extracted. At baseline and follow-up, DM was defined as 1) any occasion Hb1Ac ≥14 g/dL, 2) fasting glucose≥7 mmol/L on two occasions, or 3) with DM diagnosis. We excluded 1) patients with prior DM or with the use of antidiabetic medications; 2) patients with prior AMI/HF/AF or with the use of diuretic/beta blockers for HF were excluded. Univariate analysis and multivariate Cox analysis with adjustments on demographics, past comorbidities and medications were conducted to identify the significant risk predictors of primary and secondary outcomes. Optimal cutoffs of TyG index for the primary and secondary outcomes were found with maximally selected rank statistics approach.

**Result:** Lager TyG index is significantly associated with new onset DM (HR: 1.51, 95% CI: [1.47, 1.55], P vlaue<0.0001), new onset HF (HR: 1.27, 95% CI: [1.2, 1.34], P vlaue<0.0001), new onset AF (HR: 2.36, 95% CI: [2.26, 2.46], P vlaue<0.0001), new onset AMI (HR: 1.51, 95% CI: [1.42, 1.6], P vlaue<0.0001), new onset VTF (HR: 1.22, 95% CI: [1.13, 1.31], P vlaue<0.0001), new onset CAD (HR: 1.56, 95% CI: [1.45, 1.69], P value<0.0001) and all-cause mortality (HR: 1.21, 95% CI: [1.18, 1.25], P vlaue<0.0001). TyG index and its 3rd tertile remained significant after being adjusted with significant demographics, past comorbidities and medicactions in multivariate cox models (HR>1, P value<0.05). Optimal cut-off values of baseline TyG index and adjusted multivariate restricted cubic spline models further uncovered detailed associations of larger baseline TyG index with the primary and secondary outcome.

**Conclusion:** Higher TyG index remained significantly associated with the elevated risk of new onset DM, AF, HF, AMI, VTF, CAD and all-cause mortality after adjustments on demographics, past comorbidities, and medications.

## Introduction

The triglyceride glucose (TyG) index has been proposed to be a surrogate of insulin resistance. It has been shown to predict adverse cardiovascular events in patients with diabetes alone, or with acute coronary syndrome (1), poor prognosis in ischaemic stroke patients (2) and incident diabetes in the general population (3). In the study, we aimed to examine the relationship between TyG index and new onset DM, atrial fibrillation (AF), heart failure (HF), acute myocardial infarction (AMI), ventricular tachycardia/fibrillation (VTF), cardiovascular mortality (CAD) and all-cause mortality.

## Methods

### Study Population

The crude inclusion criteria were as follows: 29944 patients with complete triglyceride and fast glucose tests were recruited from January 1^st^, 2000 to December 31^st^, 2003 and were followed up until Dec 31^st^, 2019 (**Figure 1**). In total 5595 patients being excluded with the criteria were as follows: 1) with baseline DM diagnosis (N=1769); 2) With any prior HbA1c occasion ≥ 14 gd/L (N=1195); 3) With any two occasions of fast glucose ≥ 7 mmol/L (N=1037); 4) With prior use of antidiabetic medications (N=971); 5) With prior AMI (N=31); 6) With prior HF (N=54); 7) With prior AF (N=105); 8) With prior HF (or use of diuretics/beta blockers for HF) (N=434) before the initial date of fast glucose or triglyceride tests (baseline). A total of 24349 patients fulfilled the eligibility criteria and were included in the study cohort.

**Figure 1.**
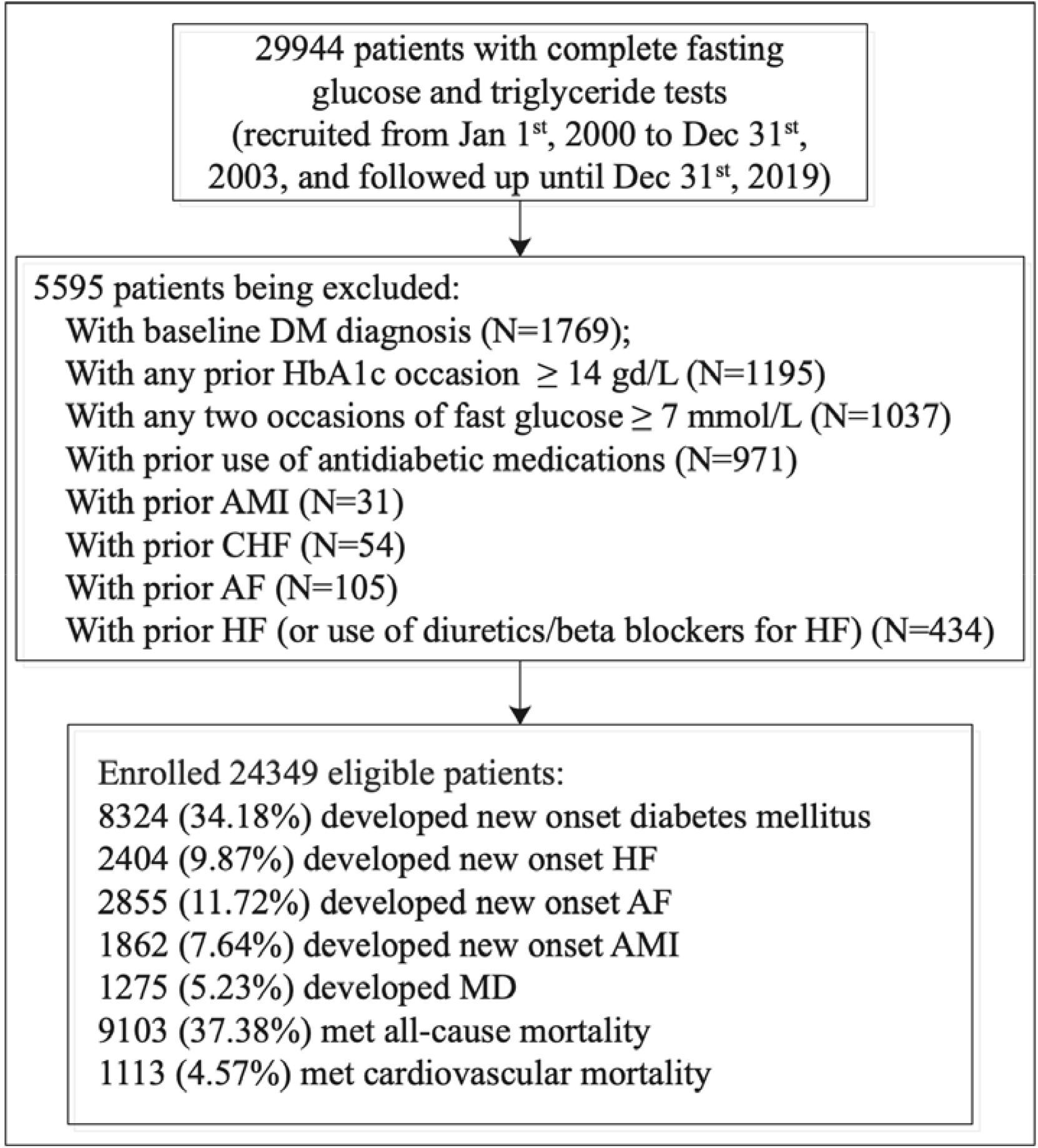
Flow chart of data processing for the study cohort.

Clinical and biochemical data were extracted for the present study. Patients’ demographics include male gender and baseline age. Prior comorbidities were extracted, including liver diseases, endocrine, hypertension, ischemic heart disease (IHD), chronic obstructive pulmonary disease (COPD), gastrointestinal, peripheral vascular disease (PVD), transient ischemic attack (TIA)/ischemic stroke, gastrointestinal bleeding, and cancer. Standard Charlson comorbidity index was also calculated. Mortality was recorded using the International Classification of Diseases Tenth Edition (ICD-10) coding, whilst the outcome of DM diagnosis and comorbidities were documented in CDARS under ICD-9 codes. ICD-10 codes I00-I78 were used to identify cardiovascular mortality. **Supplementary Table 1** displays the ICD-9 codes used to search for patient comorbidities.

Medications histories were also extracted, including angiotensinogen converting enzyme inhibitor (ACEI), angiotensin receptor blocker (ARB), calcium channel blockers, beta blockers, diuretics for HF, diuretics for hypertension, nitrates, antihypertensive drugs, anti-diabetic drugs, statins and fibrates, lipid-lowering drugs, anticoagulants, and antiplatelets. In addition, baseline biochemical data, defined as complete cell count, renal and liver function tests, and glycemic and lipid profile measured from January 1st, 2000 to December 31st, 2003 were extracted. Complete cell count include mean corpuscular volume (MCV), lymphocyte, metamyelocyte, monocyte, neutrophil, white cell count, mean corpuscular hemoglobin (MCH), myelocyte, platelet, reticulocyte, red cell count, and hematocrit (HCT). Liver and renal function tests include potassium, albumin, sodium, urea, protein, creatinine, alkaline phosphatase (ALP), aspartate transaminase, alanine aminotransferase (ALT), and bilirubin. Lipid and glycemic profile tests include low-density lipoprotein, high-density lipoprotein, HbA1c, cholesterol, fast glucose, and triglycerides. The TyG index was calculated as the ln(18×triglyceride level [mmol/L] × 18×fast glucose level [mmol/L]/2). Deciles and tertiles of TyG index were calculated.

### Primary outcomes and statistical Analysis

The primary outcome was new onset DM and the secondary outcomes included new onset AF/HF/AMI/VTF/CAD and all-cause mortality. The endpoint date of interest for all eligible patients was event presentation date, mortality, or study end (December 31, 2019). There are no records with follow-up loss. Descriptive statistics were used to summarize patients’ characteristics of the primary outcome and secondary outcomes. Continuous variables were presented as median (95% confidence interval [CI] or interquartile range [IQR]) and categorical variables were presented as count (%). The Mann-Whitney U test was used to compare two continuous variables. The Kruskal-Wallis test was used to determine whether more than two continuous variables have a different distribution. The χ2 test with Yates’ correction was used for 2×2 contingency data. The study cohort was compared according to the tertile subgroups of TyG index (**Table 1**) and those with/without event presentation (**Supplementary Tables 2-4**).

**Table 1.**
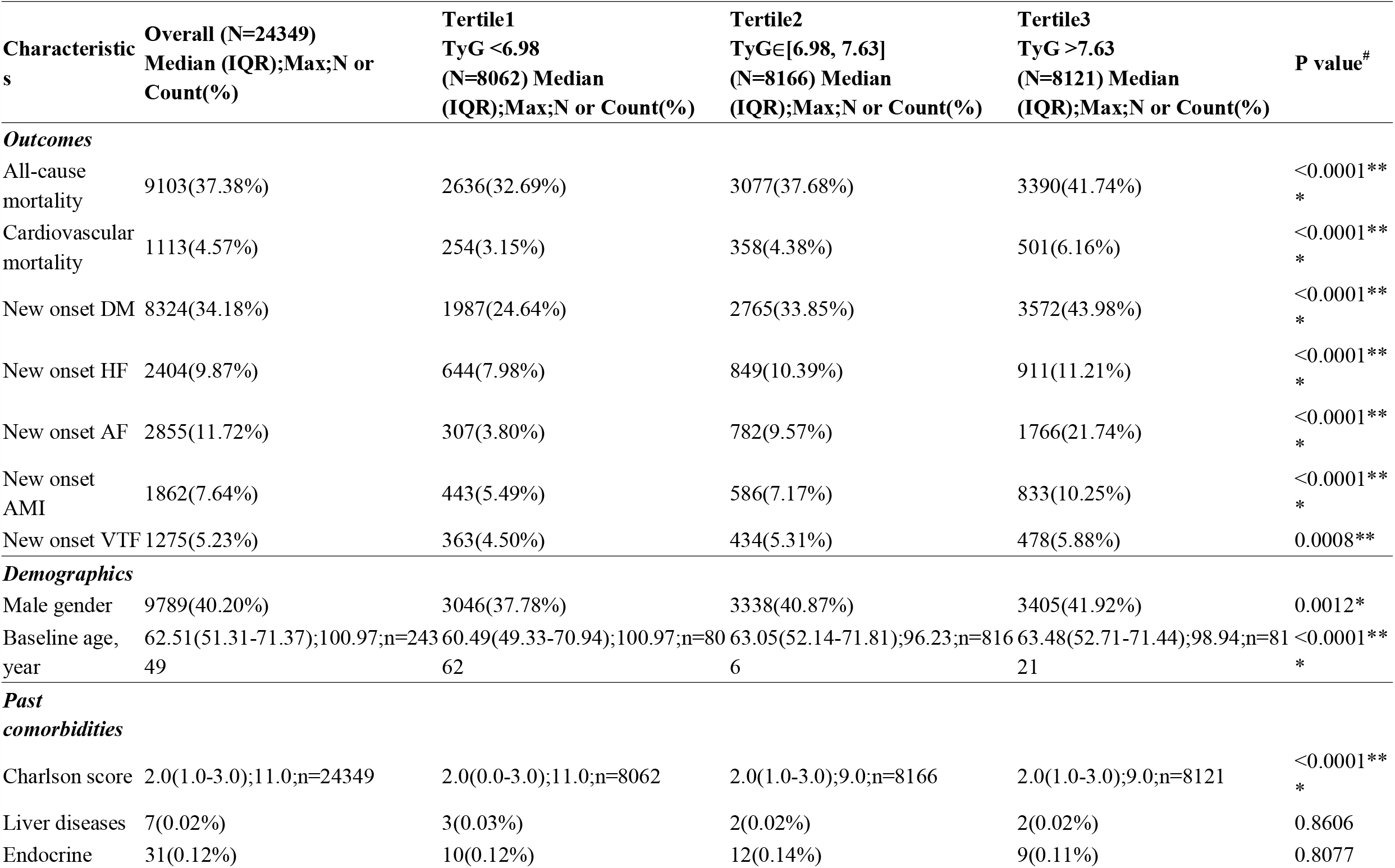
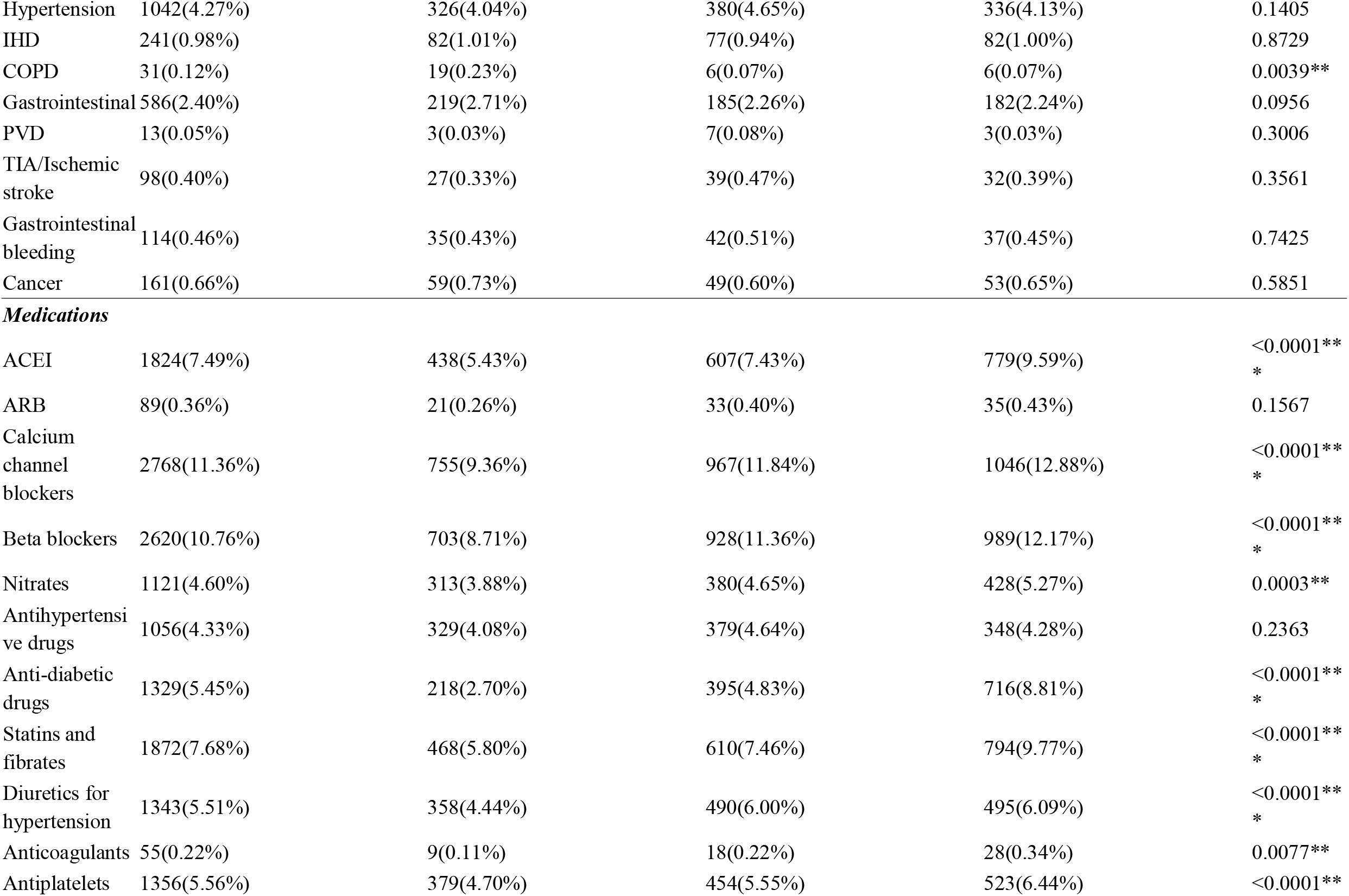
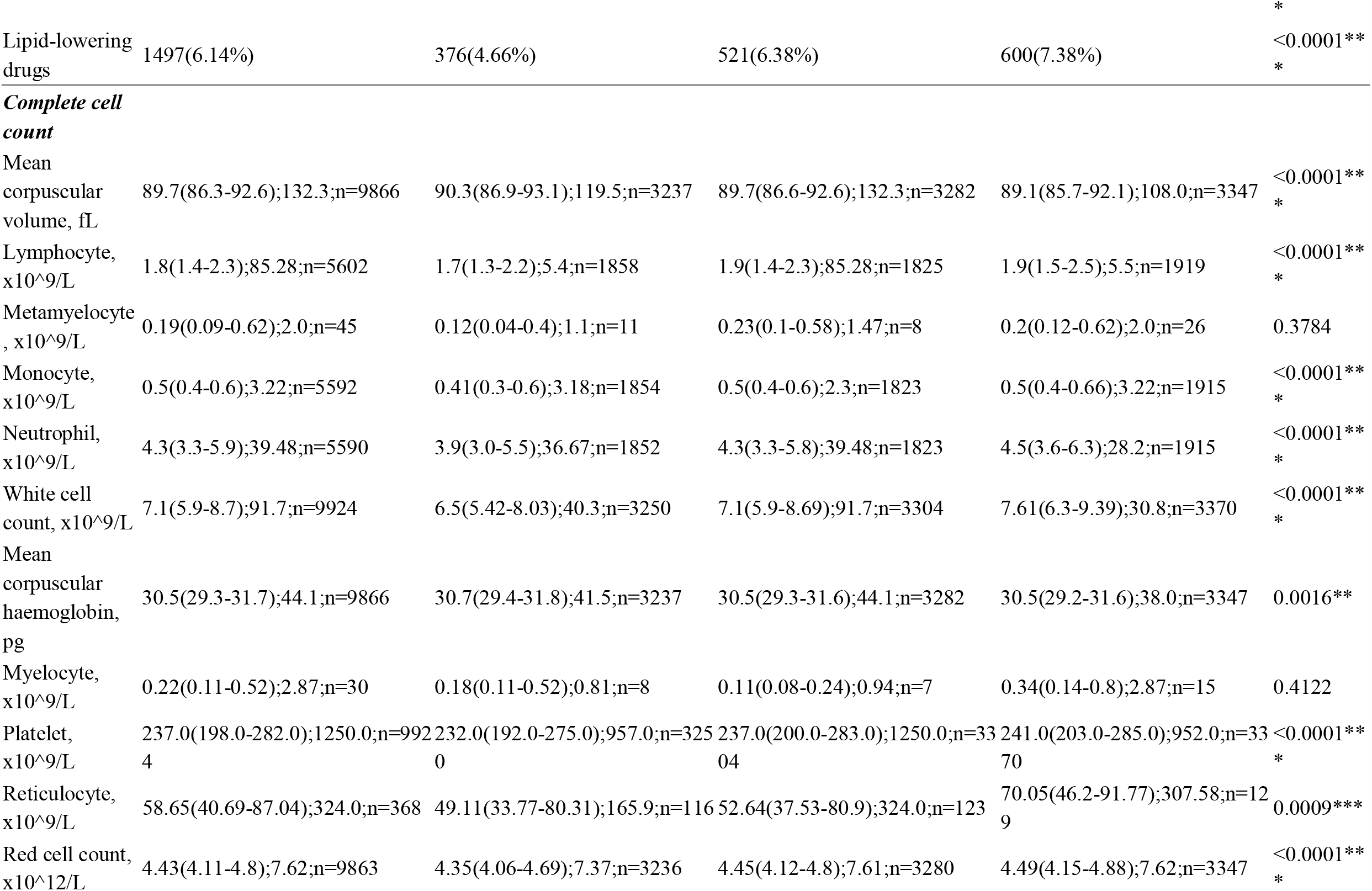
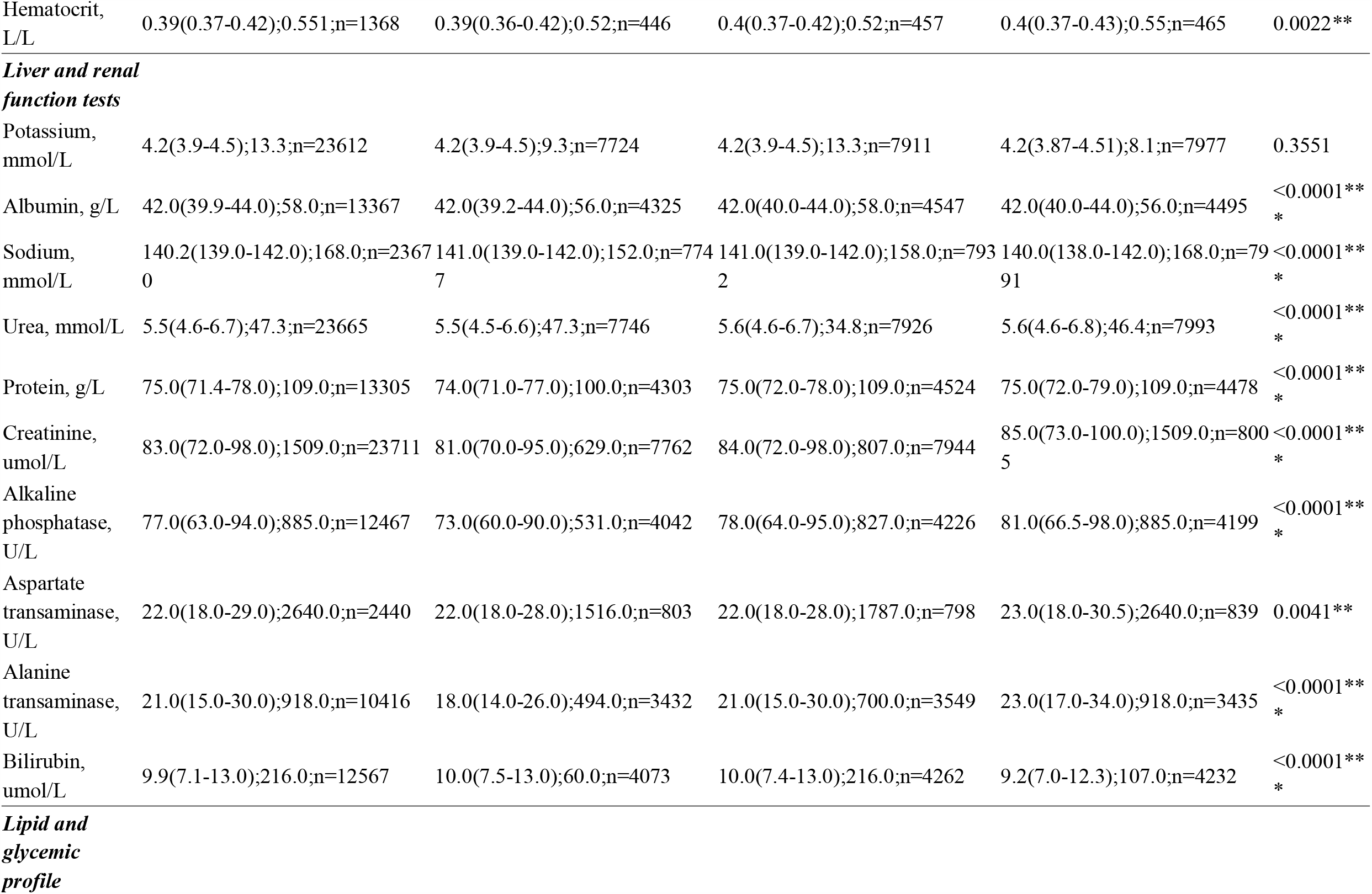
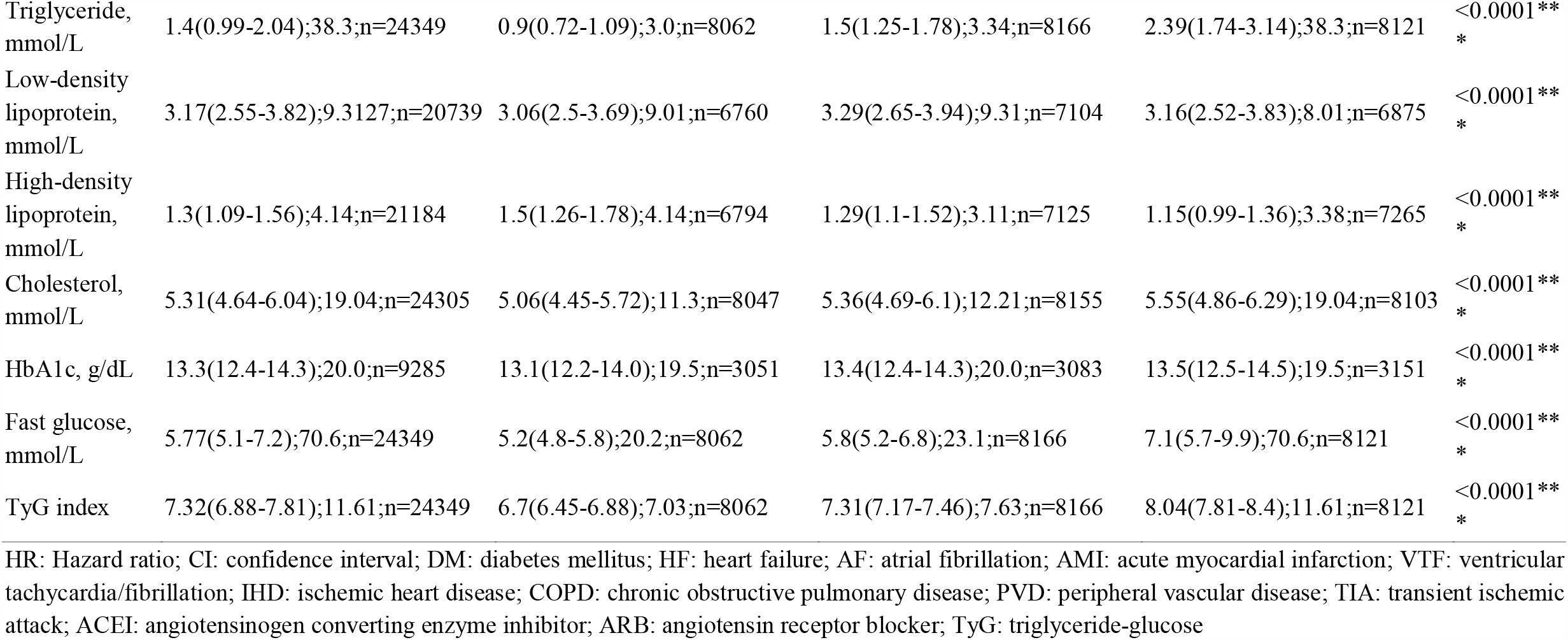
Baseline characteristics of patients according to tertile subgroups of TyG index. * for p≤ 0.05, ** for p ≤ 0.01, *** for p ≤ 0.001; # indicates the difference among three tertile subgroups

**Table 2.**
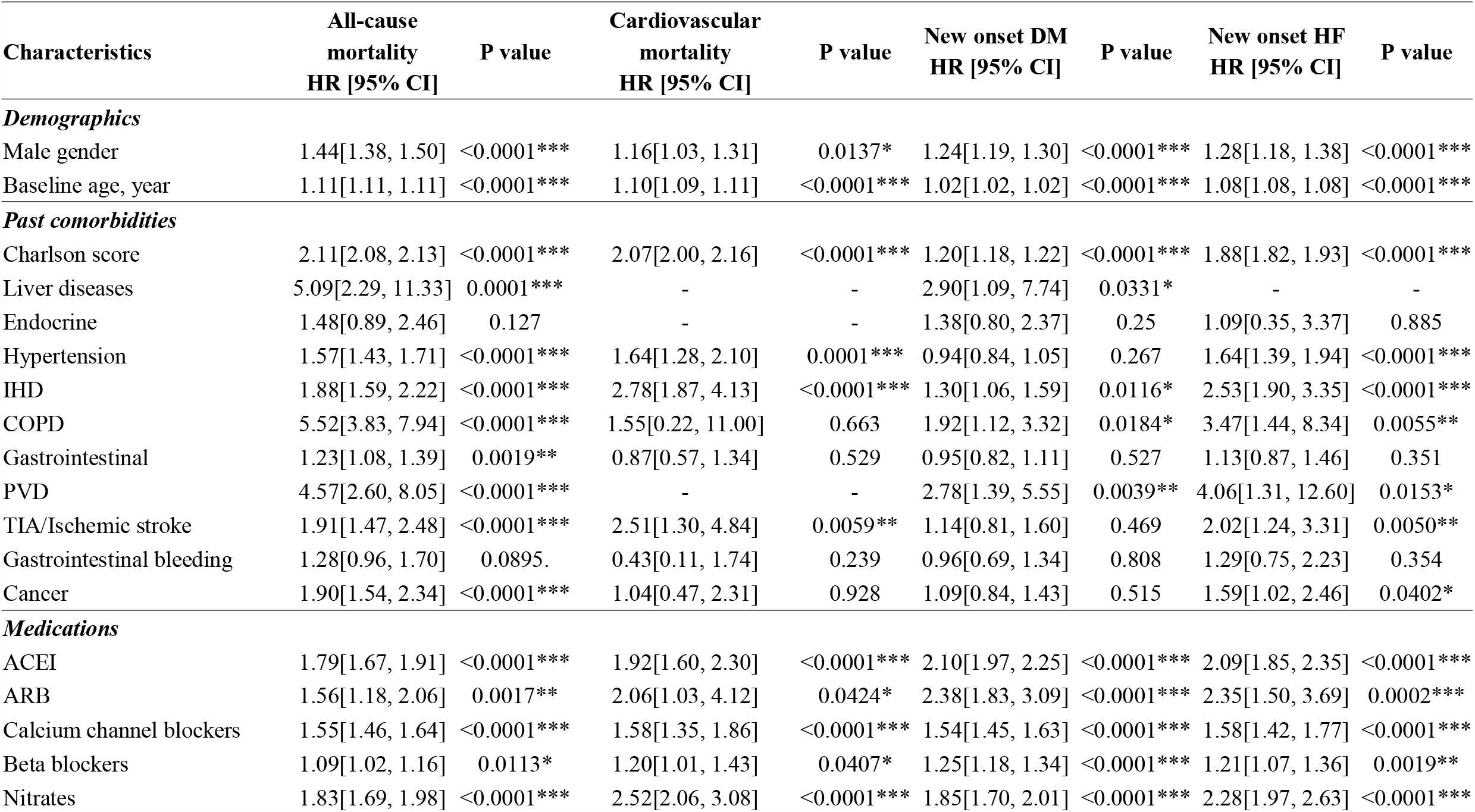
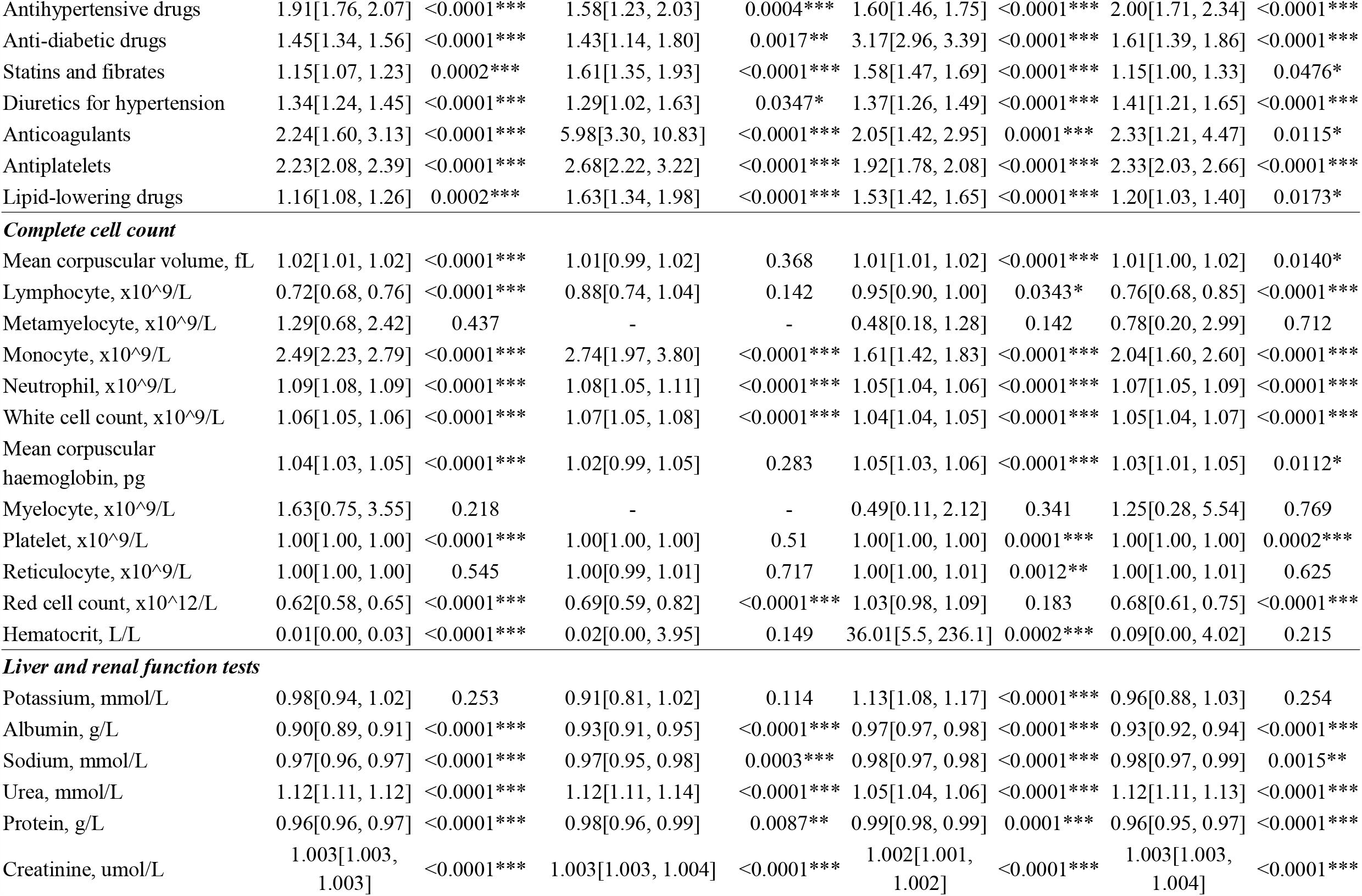
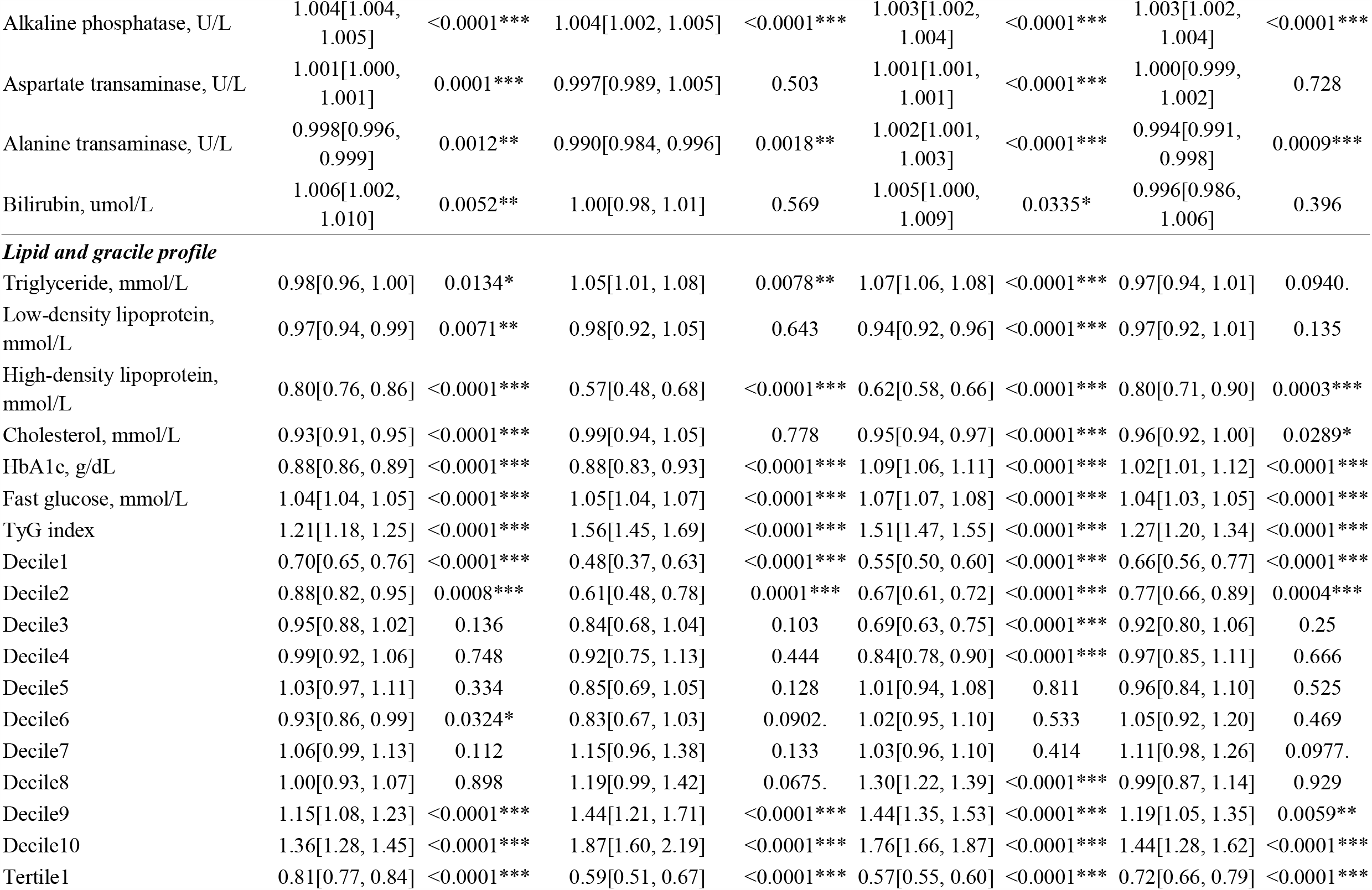
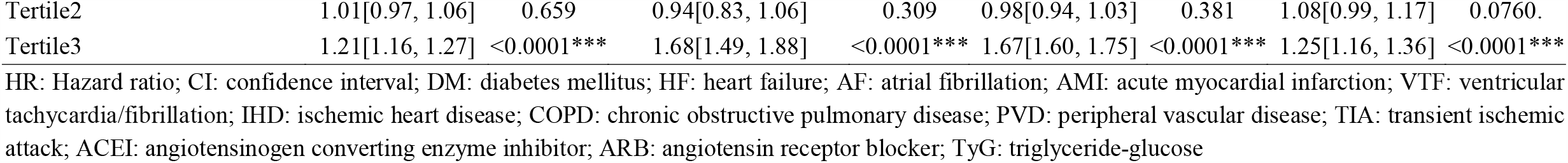
Cox-proportional hazard model analysis for all-cause mortality, cardiovascular mortality, new onset DM and new onset HF. * for p≤ 0.05, ** for p ≤ 0.01, *** for p ≤ 0.001

| Characteristics | All-cause<br>mortality<br>HR [95% CI] | P value | Cardiovascular<br>mortality<br>HR [95% CI] | P value | New onset DM<br>HR [95% CI] | P value | New onset HF<br>HR [95% CI] | P value |
| --- | --- | --- | --- | --- | --- | --- | --- | --- |
| ***Demographics*** |  |  |  |  |  |  |  |  |
| Male gender | 1.44[1.38, 1.50] | <0.0001*** | 1.16[1.03, 1.31] | 0.0137* | 1.24[1.19, 1.30] | <0.0001*** | 1.28[1.18, 1.38] | <0.0001*** |
| Baseline age, year | 1.11[1.11, 1.11] | <0.0001*** | 1.10[1.09, 1.11] | <0.0001*** | 1.02[1.02, 1.02] | <0.0001*** | 1.08[1.08, 1.08] | <0.0001*** |
| ***Past comorbidities*** |  |  |  |  |  |  |  |  |
| Charlson score | 2.11[2.08, 2.13] | <0.0001*** | 2.07[2.00, 2.16] | <0.0001*** | 1.20[1.18, 1.22] | <0.0001*** | 1.88[1.82, 1.93] | <0.0001*** |
| Liver diseases | 5.09[2.29, 11.33] | 0.0001*** | - | - | 2.90[1.09, 7.74] | 0.0331* | - | - |
| Endocrine | 1.48[0.89, 2.46] | 0.127 | - | - | 1.38[0.80, 2.37] | 0.25 | 1.09[0.35, 3.37] | 0.885 |
| Hypertension | 1.57[1.43, 1.71] | <0.0001*** | 1.64[1.28, 2.10] | 0.0001*** | 0.94[0.84, 1.05] | 0.267 | 1.64[1.39, 1.94] | <0.0001*** |
| IHD | 1.88[1.59, 2.22] | <0.0001*** | 2.78[1.87, 4.13] | <0.0001*** | 1.30[1.06, 1.59] | 0.0116* | 2.53[1.90, 3.35] | <0.0001*** |
| COPD | 5.52[3.83, 7.94] | <0.0001*** | 1.55[0.22, 11.00] | 0.663 | 1.92[1.12, 3.32] | 0.0184* | 3.47[1.44, 8.34] | 0.0055** |
| Gastrointestinal | 1.23[1.08, 1.39] | 0.0019** | 0.87[0.57, 1.34] | 0.529 | 0.95[0.82, 1.11] | 0.527 | 1.13[0.87, 1.46] | 0.351 |
| PVD | 4.57[2.60, 8.05] | <0.0001*** | - | - | 2.78[1.39, 5.55] | 0.0039** | 4.06[1.31, 12.60] | 0.0153* |
| TIA/Ischemic stroke | 1.91[1.47, 2.48] | <0.0001*** | 2.51[1.30, 4.84] | 0.0059** | 1.14[0.81, 1.60] | 0.469 | 2.02[1.24, 3.31] | 0.0050** |
| Gastrointestinal bleeding | 1.28[0.96, 1.70] | 0.0895. | 0.43[0.11, 1.74] | 0.239 | 0.96[0.69, 1.34] | 0.808 | 1.29[0.75, 2.23] | 0.354 |
| Cancer | 1.90[1.54, 2.34] | <0.0001*** | 1.04[0.47, 2.31] | 0.928 | 1.09[0.84, 1.43] | 0.515 | 1.59[1.02, 2.46] | 0.0402* |
| ***Medications*** |  |  |  |  |  |  |  |  |
| ACEI | 1.79[1.67, 1.91] | <0.0001*** | 1.92[1.60, 2.30] | <0.0001*** | 2.10[1.97, 2.25] | <0.0001*** | 2.09[1.85, 2.35] | <0.0001*** |
| ARB | 1.56[1.18, 2.06] | 0.0017** | 2.06[1.03, 4.12] | 0.0424* | 2.38[1.83, 3.09] | <0.0001*** | 2.35[1.50, 3.69] | 0.0002*** |
| Calcium channel blockers | 1.55[1.46, 1.64] | <0.0001*** | 1.58[1.35, 1.86] | <0.0001*** | 1.54[1.45, 1.63] | <0.0001*** | 1.58[1.42, 1.77] | <0.0001*** |
| Beta blockers | 1.09[1.02, 1.16] | 0.0113* | 1.20[1.01, 1.43] | 0.0407* | 1.25[1.18, 1.34] | <0.0001*** | 1.21[1.07, 1.36] | 0.0019** |
| Nitrates | 1.83[1.69, 1.98] | <0.0001*** | 2.52[2.06, 3.08] | <0.0001*** | 1.85[1.70, 2.01] | <0.0001*** | 2.28[1.97, 2.63] | <0.0001*** |
| Antihypertensive drugs | 1.91[1.76, 2.07] | <0.0001*** | 1.58[1.23, 2.03] | 0.0004*** | 1.60[1.46, 1.75] | <0.0001*** | 2.00[1.71, 2.34] | <0.0001*** |
| Anti-diabetic drugs | 1.45[1.34, 1.56] | <0.0001*** | 1.43[1.14, 1.80] | 0.0017** | 3.17[2.96, 3.39] | <0.0001*** | 1.61[1.39, 1.86] | <0.0001*** |
| Statins and fibrates | 1.15[1.07, 1.23] | 0.0002*** | 1.61[1.35, 1.93] | <0.0001*** | 1.58[1.47, 1.69] | <0.0001*** | 1.15[1.00, 1.33] | 0.0476* |
| Diuretics for hypertension | 1.34[1.24, 1.45] | <0.0001*** | 1.29[1.02, 1.63] | 0.0347* | 1.37[1.26, 1.49] | <0.0001*** | 1.41[1.21, 1.65] | <0.0001*** |
| Anticoagulants | 2.24[1.60, 3.13] | <0.0001*** | 5.98[3.30, 10.83] | <0.0001*** | 2.05[1.42, 2.95] | 0.0001*** | 2.33[1.21, 4.47] | 0.0115* |
| Antiplatelets | 2.23[2.08, 2.39] | <0.0001*** | 2.68[2.22, 3.22] | <0.0001*** | 1.92[1.78, 2.08] | <0.0001*** | 2.33[2.03, 2.66] | <0.0001*** |
| Lipid-lowering drugs | 1.16[1.08, 1.26] | 0.0002*** | 1.63[1.34, 1.98] | <0.0001*** | 1.53[1.42, 1.65] | <0.0001*** | 1.20[1.03, 1.40] | 0.0173* |
| ***Complete cell count*** |  |  |  |  |  |  |  |  |
| Mean corpuscular volume, fL | 1.02[1.01, 1.02] | <0.0001*** | 1.01[0.99, 1.02] | 0.368 | 1.01[1.01, 1.02] | <0.0001*** | 1.01[1.00, 1.02] | 0.0140* |
| Lymphocyte, x10<sup>9</sup>/L | 0.72[0.68, 0.76] | <0.0001*** | 0.88[0.74, 1.04] | 0.142 | 0.95[0.90, 1.00] | 0.0343* | 0.76[0.68, 0.85] | <0.0001*** |
| Metamyelocyte, x10<sup>9</sup>/L | 1.29[0.68, 2.42] | 0.437 | - | - | 0.48[0.18, 1.28] | 0.142 | 0.78[0.20, 2.99] | 0.712 |
| Monocyte, x10<sup>9</sup>/L | 2.49[2.23, 2.79] | <0.0001*** | 2.74[1.97, 3.80] | <0.0001*** | 1.61[1.42, 1.83] | <0.0001*** | 2.04[1.60, 2.60] | <0.0001*** |
| Neutrophil, x10<sup>9</sup>/L | 1.09[1.08, 1.09] | <0.0001*** | 1.08[1.05, 1.11] | <0.0001*** | 1.05[1.04, 1.06] | <0.0001*** | 1.07[1.05, 1.09] | <0.0001*** |
| White cell count, x10<sup>9</sup>/L | 1.06[1.05, 1.06] | <0.0001*** | 1.07[1.05, 1.08] | <0.0001*** | 1.04[1.04, 1.05] | <0.0001*** | 1.05[1.04, 1.07] | <0.0001*** |
| Mean corpuscular haemoglobin, pg | 1.04[1.03, 1.05] | <0.0001*** | 1.02[0.99, 1.05] | 0.283 | 1.05[1.03, 1.06] | <0.0001*** | 1.03[1.01, 1.05] | 0.0112* |
| Myelocyte, x10<sup>9</sup>/L | 1.63[0.75, 3.55] | 0.218 | - | - | 0.49[0.11, 2.12] | 0.341 | 1.25[0.28, 5.54] | 0.769 |
| Platelet, x10<sup>9</sup>/L | 1.00[1.00, 1.00] | <0.0001*** | 1.00[1.00, 1.00] | 0.51 | 1.00[1.00, 1.00] | 0.0001*** | 1.00[1.00, 1.00] | 0.0002*** |
| Reticulocyte, x10<sup>9</sup>/L | 1.00[1.00, 1.00] | 0.545 | 1.00[0.99, 1.01] | 0.717 | 1.00[1.00, 1.01] | 0.0012** | 1.00[1.00, 1.01] | 0.625 |
| Red cell count, x10<sup>12</sup>/L | 0.62[0.58, 0.65] | <0.0001*** | 0.69[0.59, 0.82] | <0.0001*** | 1.03[0.98, 1.09] | 0.183 | 0.68[0.61, 0.75] | <0.0001*** |
| Hematocrit, L/L | 0.01[0.00, 0.03] | <0.0001*** | 0.02[0.00, 3.95] | 0.149 | 36.01[5.5, 236.1] | 0.0002*** | 0.09[0.00, 4.02] | 0.215 |
| ***Liver and renal function tests*** |  |  |  |  |  |  |  |  |
| Potassium, mmol/L | 0.98[0.94, 1.02] | 0.253 | 0.91[0.81, 1.02] | 0.114 | 1.13[1.08, 1.17] | <0.0001*** | 0.96[0.88, 1.03] | 0.254 |
| Albumin, g/L | 0.90[0.89, 0.91] | <0.0001*** | 0.93[0.91, 0.95] | <0.0001*** | 0.97[0.97, 0.98] | <0.0001*** | 0.93[0.92, 0.94] | <0.0001*** |
| Sodium, mmol/L | 0.97[0.96, 0.97] | <0.0001*** | 0.97[0.95, 0.98] | 0.0003*** | 0.98[0.97, 0.98] | <0.0001*** | 0.98[0.97, 0.99] | 0.0015** |
| Urea, mmol/L | 1.12[1.11, 1.12] | <0.0001*** | 1.12[1.11, 1.14] | <0.0001*** | 1.05[1.04, 1.06] | <0.0001*** | 1.12[1.11, 1.13] | <0.0001*** |
| Protein, g/L | 0.96[0.96, 0.97] | <0.0001*** | 0.98[0.96, 0.99] | 0.0087** | 0.99[0.98, 0.99] | 0.0001*** | 0.96[0.95, 0.97] | <0.0001*** |
| Creatinine, umol/L | 1.003[1.003, 1.003] | <0.0001*** | 1.003[1.003, 1.004] | <0.0001*** | 1.002[1.001, 1.002] | <0.0001*** | 1.003[1.003, 1.004] | <0.0001*** |
| Alkaline phosphatase, U/L | 1.004[1.004, 1.005] | <0.0001*** | 1.004[1.002, 1.005] | <0.0001*** | 1.003[1.002, 1.004] | <0.0001*** | 1.003[1.002, 1.004] | <0.0001*** |
| Aspartate transaminase, U/L | 1.001[1.000, 1.001] | 0.0001*** | 0.997[0.989, 1.005] | 0.503 | 1.001[1.001, 1.001] | <0.0001*** | 1.000[0.999, 1.002] | 0.728 |
| Alanine transaminase, U/L | 0.998[0.996, 0.999] | 0.0012** | 0.990[0.984, 0.996] | 0.0018** | 1.002[1.001, 1.003] | <0.0001*** | 0.994[0.991, 0.998] | 0.0009*** |
| Bilirubin, umol/L | 1.006[1.002, 1.010] | 0.0052** | 1.00[0.98, 1.01] | 0.569 | 1.005[1.000, 1.009] | 0.0335* | 0.996[0.986, 1.006] | 0.396 |
| ***Lipid and gracile profile*** |  |  |  |  |  |  |  |  |
| Triglyceride, mmol/L | 0.98[0.96, 1.00] | 0.0134* | 1.05[1.01, 1.08] | 0.0078** | 1.07[1.06, 1.08] | <0.0001*** | 0.97[0.94, 1.01] | 0.0940. |
| Low-density lipoprotein, mmol/L | 0.97[0.94, 0.99] | 0.0071** | 0.98[0.92, 1.05] | 0.643 | 0.94[0.92, 0.96] | <0.0001*** | 0.97[0.92, 1.01] | 0.135 |
| High-density lipoprotein, mmol/L | 0.80[0.76, 0.86] | <0.0001*** | 0.57[0.48, 0.68] | <0.0001*** | 0.62[0.58, 0.66] | <0.0001*** | 0.80[0.71, 0.90] | 0.0003*** |
| Cholesterol, mmol/L | 0.93[0.91, 0.95] | <0.0001*** | 0.99[0.94, 1.05] | 0.778 | 0.95[0.94, 0.97] | <0.0001*** | 0.96[0.92, 1.00] | 0.0289* |
| HbA1c, g/dL | 0.88[0.86, 0.89] | <0.0001*** | 0.88[0.83, 0.93] | <0.0001*** | 1.09[1.06, 1.11] | <0.0001*** | 1.02[1.01, 1.12] | <0.0001*** |
| Fast glucose, mmol/L | 1.04[1.04, 1.05] | <0.0001*** | 1.05[1.04, 1.07] | <0.0001*** | 1.07[1.07, 1.08] | <0.0001*** | 1.04[1.03, 1.05] | <0.0001*** |
| TyG index | 1.21[1.18, 1.25] | <0.0001*** | 1.56[1.45, 1.69] | <0.0001*** | 1.51[1.47, 1.55] | <0.0001*** | 1.27[1.20, 1.34] | <0.0001*** |
| Decile1 | 0.70[0.65, 0.76] | <0.0001*** | 0.48[0.37, 0.63] | <0.0001*** | 0.55[0.50, 0.60] | <0.0001*** | 0.66[0.56, 0.77] | <0.0001*** |
| Decile2 | 0.88[0.82, 0.95] | 0.0008*** | 0.61[0.48, 0.78] | 0.0001*** | 0.67[0.61, 0.72] | <0.0001*** | 0.77[0.66, 0.89] | 0.0004*** |
| Decile3 | 0.95[0.88, 1.02] | 0.136 | 0.84[0.68, 1.04] | 0.103 | 0.69[0.63, 0.75] | <0.0001*** | 0.92[0.80, 1.06] | 0.25 |
| Decile4 | 0.99[0.92, 1.06] | 0.748 | 0.92[0.75, 1.13] | 0.444 | 0.84[0.78, 0.90] | <0.0001*** | 0.97[0.85, 1.11] | 0.666 |
| Decile5 | 1.03[0.97, 1.11] | 0.334 | 0.85[0.69, 1.05] | 0.128 | 1.01[0.94, 1.08] | 0.811 | 0.96[0.84, 1.10] | 0.525 |
| Decile6 | 0.93[0.86, 0.99] | 0.0324* | 0.83[0.67, 1.03] | 0.0902. | 1.02[0.95, 1.10] | 0.533 | 1.05[0.92, 1.20] | 0.469 |
| Decile7 | 1.06[0.99, 1.13] | 0.112 | 1.15[0.96, 1.38] | 0.133 | 1.03[0.96, 1.10] | 0.414 | 1.11[0.98, 1.26] | 0.0977. |
| Decile8 | 1.00[0.93, 1.07] | 0.898 | 1.19[0.99, 1.42] | 0.0675. | 1.30[1.22, 1.39] | <0.0001*** | 0.99[0.87, 1.14] | 0.929 |
| Decile9 | 1.15[1.08, 1.23] | <0.0001*** | 1.44[1.21, 1.71] | <0.0001*** | 1.44[1.35, 1.53] | <0.0001*** | 1.19[1.05, 1.35] | 0.0059** |
| Decile10 | 1.36[1.28, 1.45] | <0.0001*** | 1.87[1.60, 2.19] | <0.0001*** | 1.76[1.66, 1.87] | <0.0001*** | 1.44[1.28, 1.62] | <0.0001*** |
| Tertile1 | 0.81[0.77, 0.84] | <0.0001*** | 0.59[0.51, 0.67] | <0.0001*** | 0.57[0.55, 0.60] | <0.0001*** | 0.72[0.66, 0.79] | <0.0001*** |
| Tertile2 | 1.01[0.97, 1.06] | 0.659 | 0.94[0.83, 1.06] | 0.309 | 0.98[0.94, 1.03] | 0.381 | 1.08[0.99, 1.17] | 0.0760. |
| Tertile3 | 1.21[1.16, 1.27] | <0.0001*** | 1.68[1.49, 1.88] | <0.0001*** | 1.67[1.60, 1.75] | <0.0001*** | 1.25[1.16, 1.36] | <0.0001*** |

HR: Hazard ratio; CI: confidence interval; DM: diabetes mellitus; HF: heart failure; AF: atrial fibrillation; AMI: acute myocardial infarction; VTF: ventricular tachycardia/fibrillation; IHD: ischemic heart disease; COPD: chronic obstructive pulmonary disease; PVD: peripheral vascular disease; TIA: transient ischemic attack; ACEI: angiotensinogen converting enzyme inhibitor; ARB: angiotensin receptor blocker; TyG: triglyceride-glucose

Univariate Cox hazard proportional analysis models were used to identify the significant risk predictors of primary and secondary outcomes. Hazard ratios (HRs) with corresponding 95% CIs and P values were reported accordingly. Kaplan-Meier curves were plotted against the time-to-event for primary and secondary outcomes stratified by deciles of TyG index (**Figure 2**). Adjusted cubic spline models of the associations between TyG index and the risk of primary and secondary outcomes were presented (**Figure 3**). Hazard ratios (HRs) of primary and secondary outcomes according to tertiles of TyG index were summarized (**Table 4**). Optimal cut-offs of baseline TyG index for primary and secondary outcomes were derived with maximal survival rank statistics approach after adjusting for significant demographics, past comorbidities, and medications with multivariate Cox model, and compared with 1^st^ tertile subgroup as reference (**Table 5**). Adjusted cubic spline model of the associations between TyG index and risk of new-onset DM and all-cause mortality were presented (**Figure 4**). All significance tests were two-tailed and considered statistically significant if P values were 0.05. Data analyses were performed using RStudio software (Version: 1.1.456) and Python (Version: 3.6).

**Table 3.**
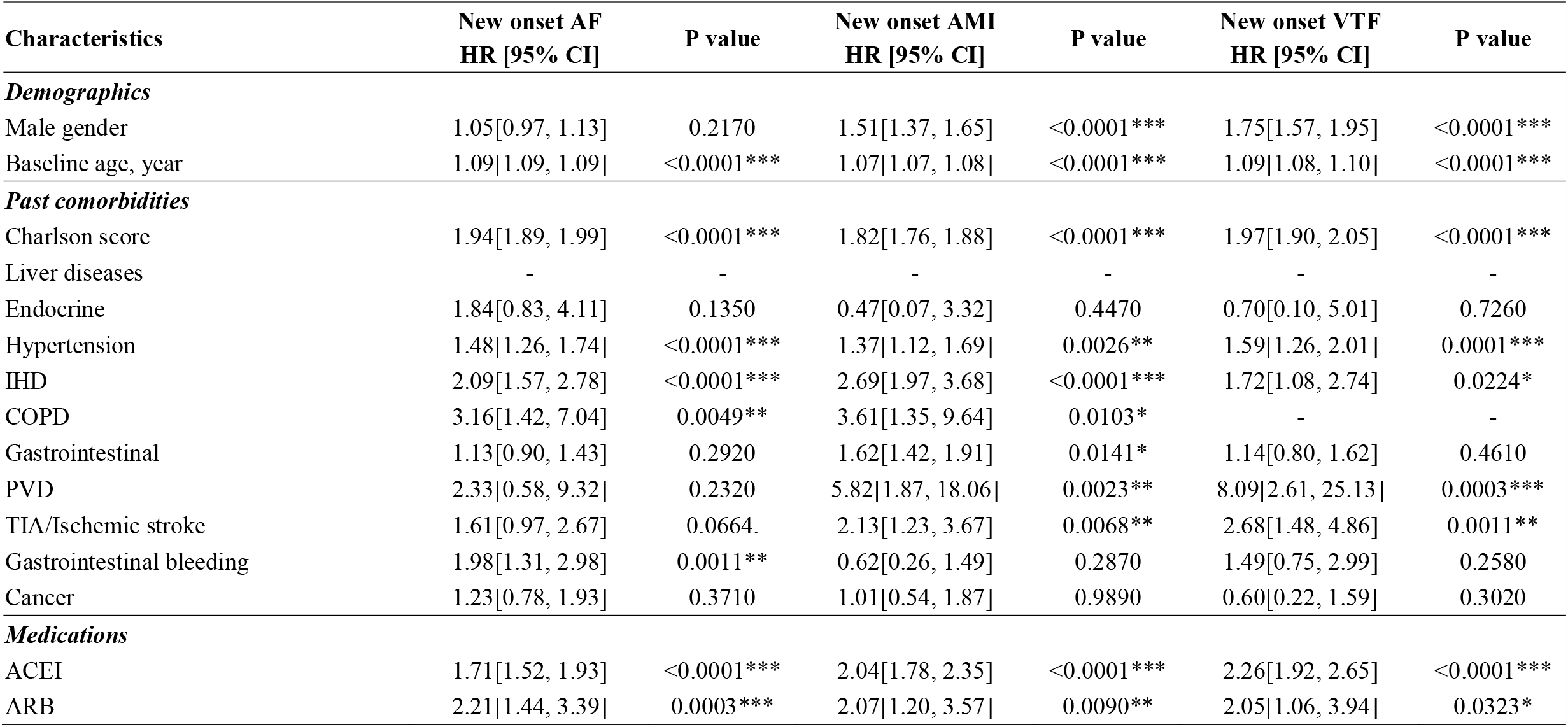
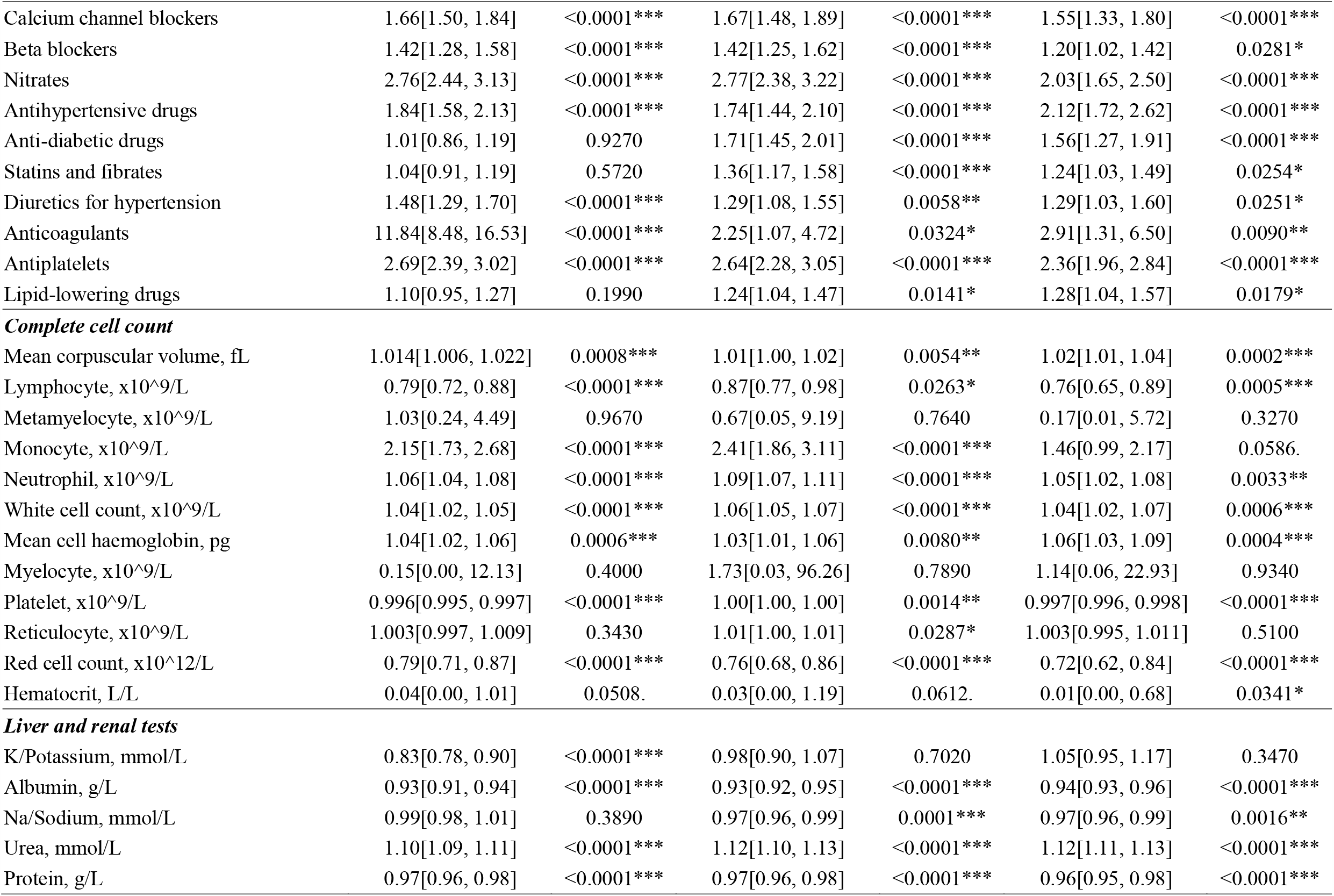
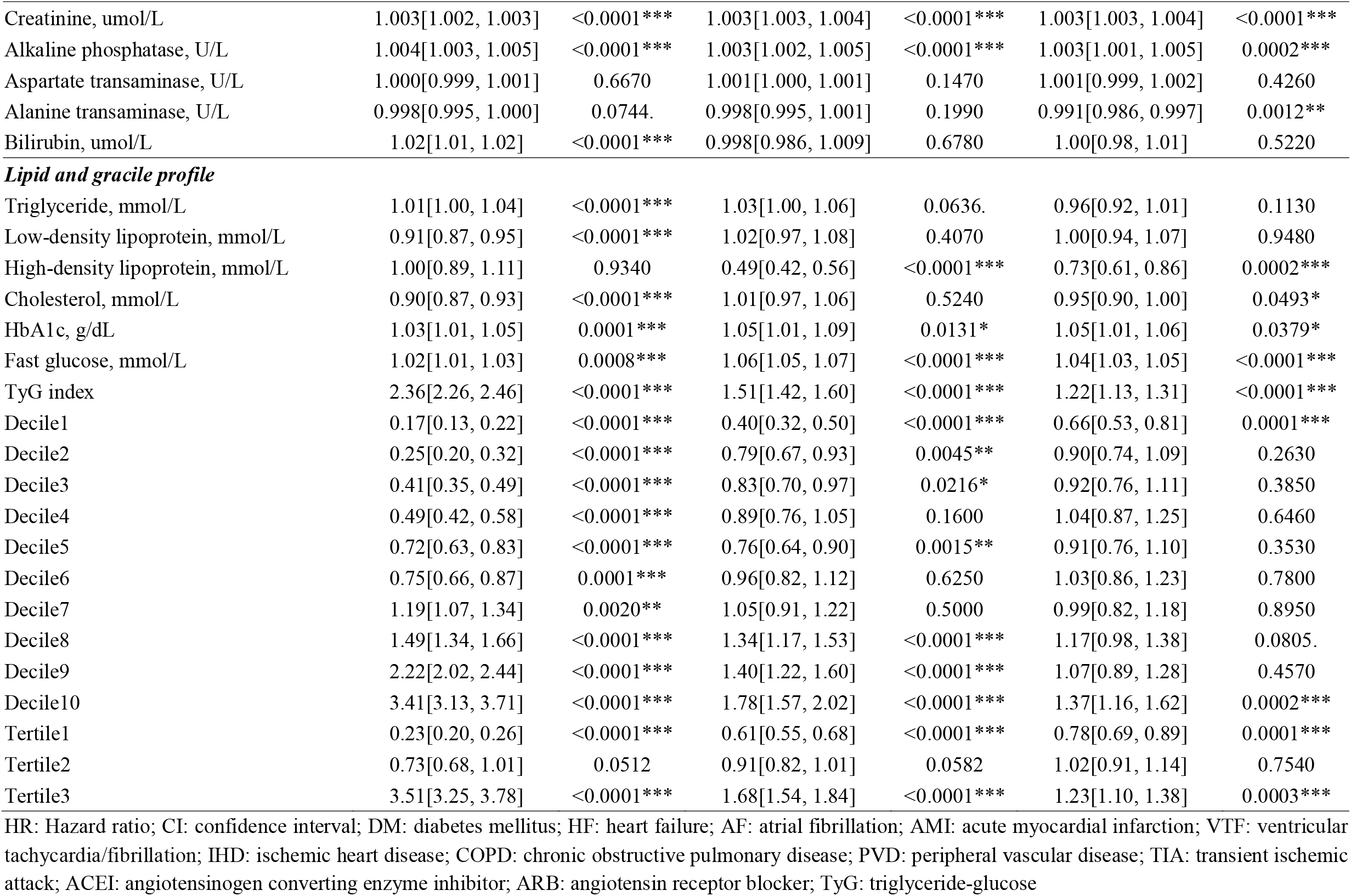
Cox-proportional hazard model analysis for new onset AF, AMI, and VTF. * for p≤ 0.05, ** for p ≤ 0.01, *** for p ≤ 0.001

**Table 4.** The hazard ratios (HRs) of primary and secondary outcomes according to tertiles of TyG index. * for p≤ 0.05, ** for p ≤ 0.01, *** for p ≤ 0.001 HR hazard ratios; IR: incidence rate. Model 1 adjusted for none. Model 2 adjusted for significant demographics. Model 3 adjusted for significant demographics and past comorbidities. Model 4 adjusted for significant demographics, past comorbidities and medications.

| Outcomes | Model 1 |  |  | Model 2 |  |  |
| --- | --- | --- | --- | --- | --- | --- |
|  | TyG <6.98 | TyG ∈ [6.98, 7.63] | TyG >7.63 | TyG <6.98 | TyG ∈ [6.98, 7.63] | TyG >7.63 |
| All-cause mortality | 1.00 (reference) | 1.01[0.97, 1.06] | 1.21[1.16, 1.27]*** | 1.00 (reference) | 0.96[0.92, 1.01] | 1.19 [1.14, 1.24]*** |
| CAD mortality | 1.00 (reference) | 0.94[0.83, 1.06] | 1.68[1.49, 1.88]*** | 1.00 (reference) | 0.89[0.78, 1.01] | 1.62[1.44, 1.82]*** |
| New onset DM | 1.00 (reference) | 0.98[0.94, 1.03] | 1.67[1.60, 1.75]*** | 1.00 (reference) | 0.96[0.92, 1.01] | 1.63[1.56, 1.70]*** |
| New onset HF | 1.00 (reference) | 1.08[0.99, 1.17] | 1.25[1.16, 1.36]*** | 1.00 (reference) | 1.03[0.95, 1.12] | 1.21[1.11, 1.31]*** |
| New onset AF | 1.00 (reference) | 0.73[0.68, 1.01] | 3.51[3.25, 3.78]*** | 1.00 (reference) | 0.7[0.64, 1.01] | 3.48[3.23, 3.76]*** |
| New onset AMI | 1.00 (reference) | 0.91[0.82, 1.01] | 1.68[1.54, 1.84]*** | 1.00 (reference) | 0.87[0.79, 1.01] | 1.62[1.48, 1.78]*** |
| New onset VTF | 1.00 (reference) | 1.02[0.91, 1.14] | 1.23[1.10, 1.38]*** | 1.00 (reference) | 0.97[0.87, 1.09] | 1.2[1.07, 1.35]*** |
| Outcomes | Model 3 |  |  | Model 4 |  |  |
|  | TyG <6.98 | TyG ∈ [6.98, 7.63] | TyG >7.63 | TyG <6.98 | TyG ∈ [6.98, 7.63] | TyG >7.63 |
| All-cause mortality | 1.00 (reference) | 0.96 [0.92, 1.01] | 1.19 [1.14, 1.24]*** | 1.00 (reference) | 0.96[0.92, 1.01] | 1.17 [1.12, 1.22]*** |
| CAD mortality | 1.00 (reference) | 0.89[0.79, 1.01] | 1.62[1.44, 1.82]*** | 1.00 (reference) | 0.89[0.79,1.03] | 1.59[1.41, 1.79]*** |
| New onset DM | 1.00 (reference) | 0.96[0.92, 1.01] | 1.63[1.56, 1.7]*** | 1.00 (reference) | 0.98[0.93, 1.02] | 1.52[1.46, 1.59]*** |
| New onset HF | 1.00 (reference) | 1.03[0.95, 1.12] | 1.21[1.12, 1.31]*** | 1.00 (reference) | 1.03[0.95, 1.12] | 1.19[1.09, 1.29]*** |
| New onset AF | 1.00 (reference) | 0.7[0.64, 1.01] | 3.49[3.23, 3.76]*** | 1.00 (reference) | 0.68[0.63 ,1.04] | 3.61[3.34, 3.89]*** |
| New onset AMI | 1.00 (reference) | 0.87[0.79, 1.01] | 1.63[1.48, 1.78]*** | 1.00 (reference) | 0.87[0.79, 1.06] | 1.59[1.45, 1.74]*** |
| New onset VTF | 1.00 (reference) | 0.97[0.86, 1.09] | 1.2[1.08, 1.35]** | 1.00 (reference) | 0.97[0.86, 1.09] | 1.18[1.06, 1.33]** |

HR: Hazard ratio; CI: confidence interval; DM: diabetes mellitus; HF: heart failure; AF: atrial fibrillation; AMI: acute myocardial infarction; VTF: ventricular tachycardia/fibrillation; TyG: triglyceride-glucose

**Table 5.** Optimal cut-offs between triglyceride level and primary and secondary outcomes with maximal survival rank statistics approach. * for p≤ 0.05, ** for p ≤ 0.01, *** for p ≤ 0.001

|  | **TyG cutoff value** | **<Cut-off value**<br>**HR [95% CI]** | **≥Cut-off value**<br>**HR [95% CI]** | **P for log ratio test** |
| --- | --- | --- | --- | --- |
| All-cause mortality | 8.05 | 1.0 (reference) | 1.34[1.28, 1.42]*** | <0.0001 |
| Cardiovascular mortality | 7.43 | 1.0 (reference) | 1.79[1.59, 2.02]*** | <0.0001 |
| New onset DM | 7.68 | 1.0 (reference) | 1.71[1.64, 1.79]*** | <0.0001 |
| New onset HF | 7.24 | 1.0 (reference) | 1.35[1.24, 1.47]*** | <0.0001 |
| New onset AF | 7.75 | 1.0 (reference) | 3.58[3.33, 3.86]*** | <0.0001 |
| New onset AMI | 7.66 | 1.0 (reference) | 1.75[1.6, 1.92]*** | <0.0001 |
| New onset VTF | 7.79 | 1.0 (reference) | 1.33[1.18, 1.5]*** | <0.0001 |

HR: Hazard ratio; CI: confidence interval; DM: diabetes mellitus; HF: heart failure; AF: atrial fibrillation; AMI: acute myocardial infarction; VTF: ventricular tachycardia/fibrillation; TyG: triglyceride-glucose
Multivariate Cox regression was performed to examine the association between triglyceride levels and primary and secondary outcomes; Effects include significant demographics, past comorbidities, and medications.

**Figure 2.**
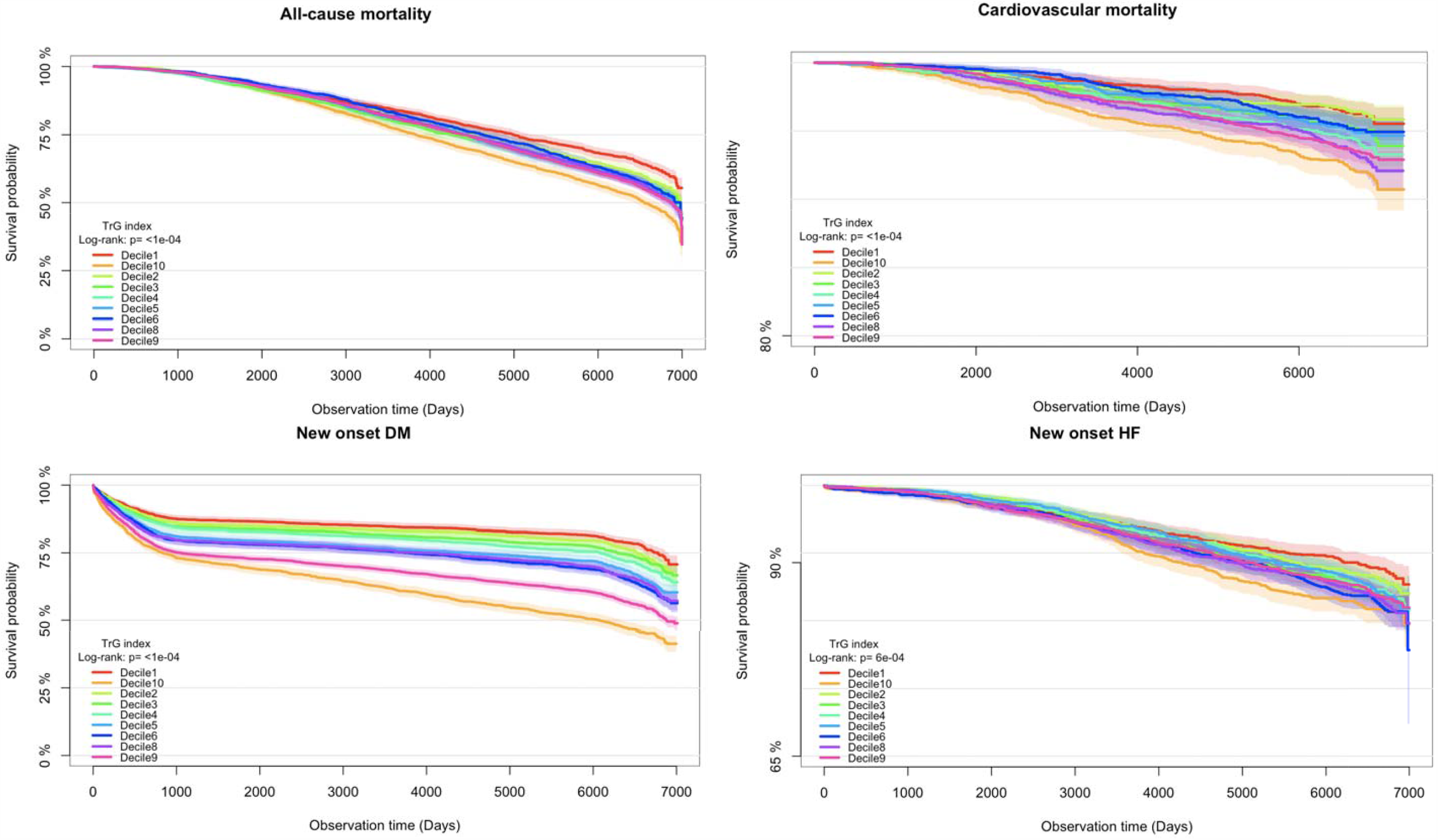
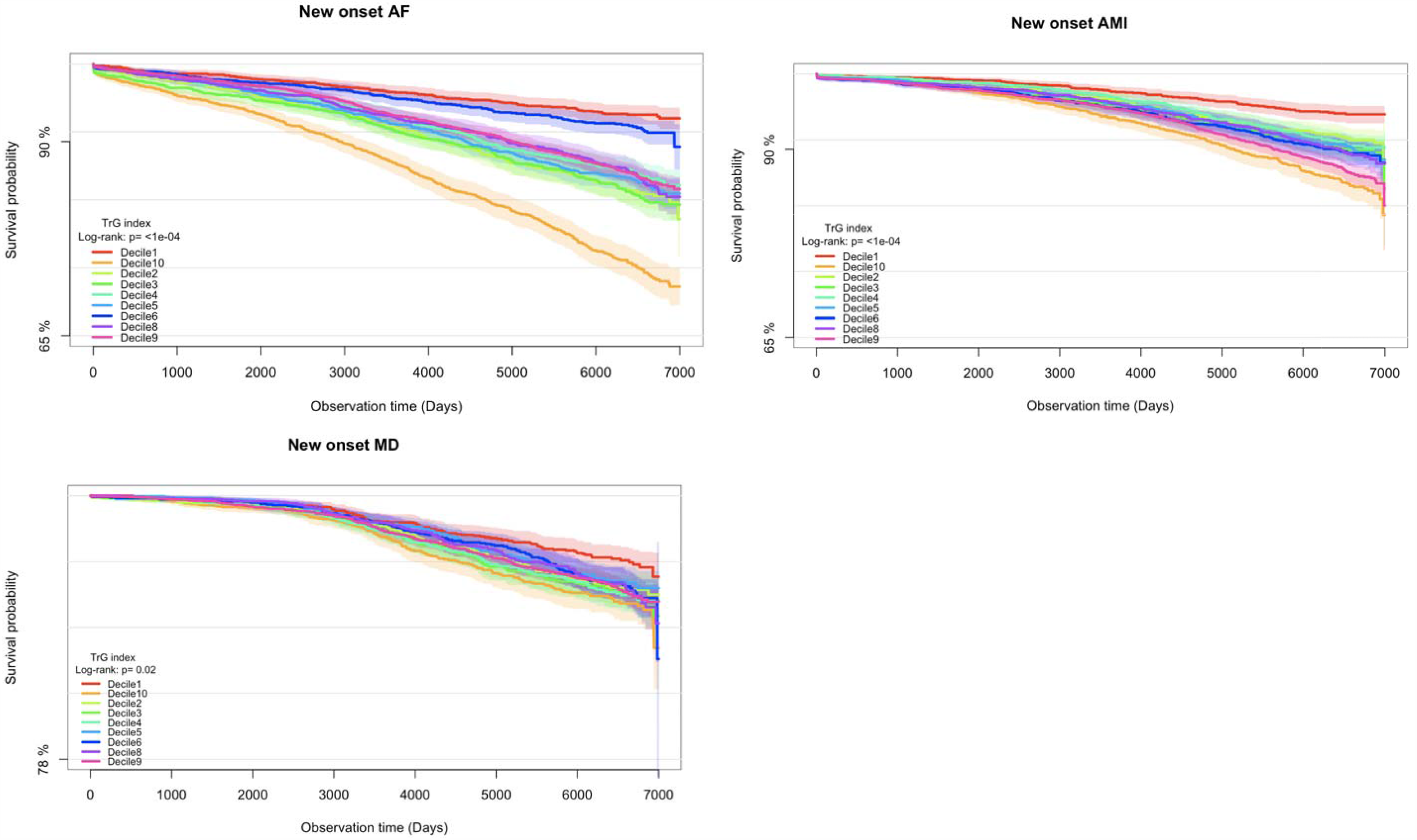
Kaplan-Meier survival curves for primary and secondary outcomes stratified by TyG index deciles. TyG: triglyceride glucose.

**Figure 3.**
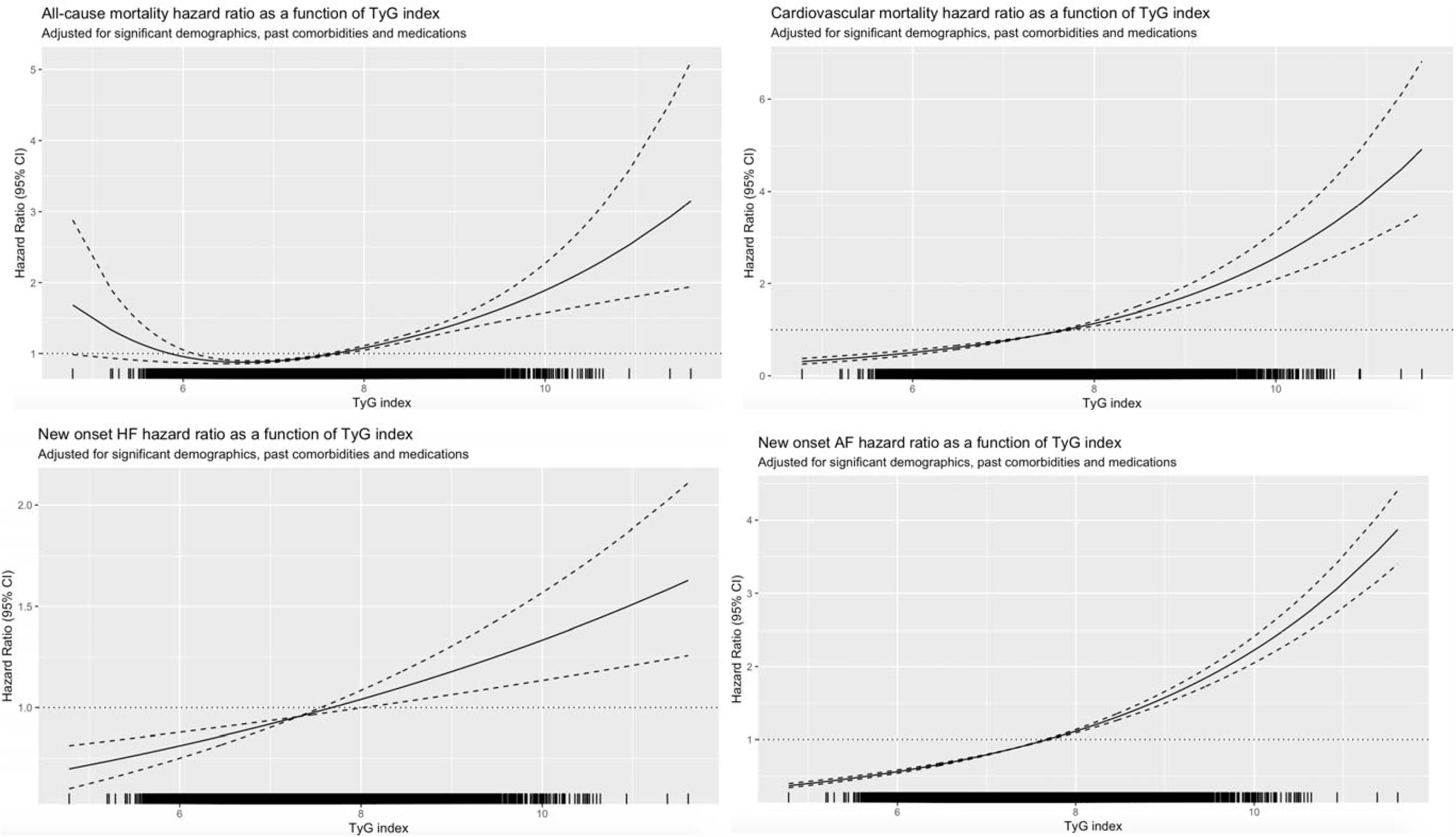
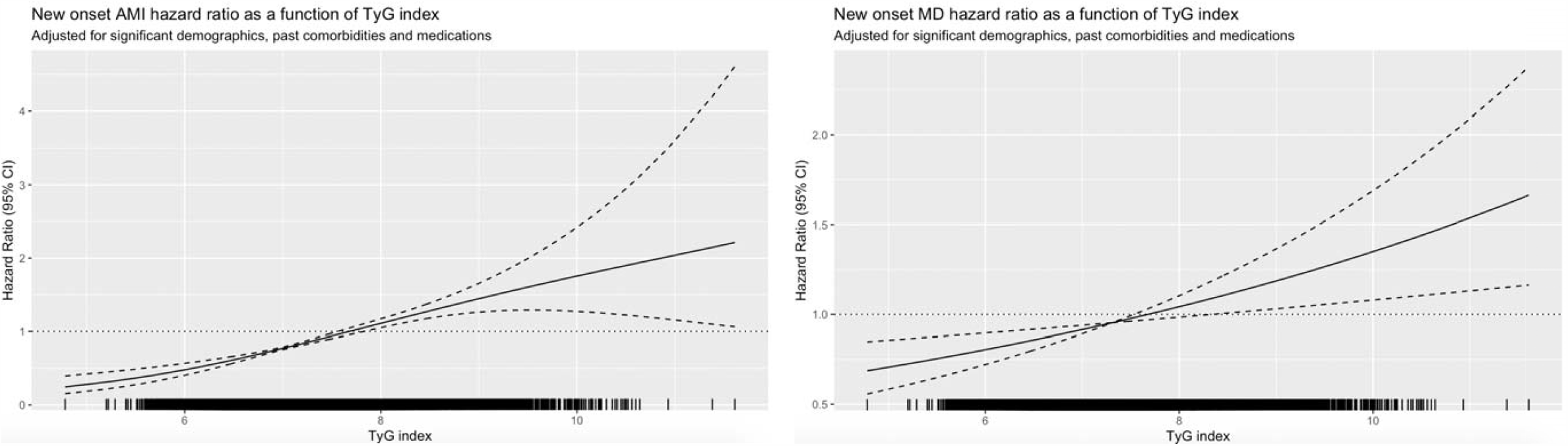
Adjusted cubic spline model of the associations between TyG index and risk of primary and secondary outcomes. TyG: triglyceride gluco.

**Figure 4.**
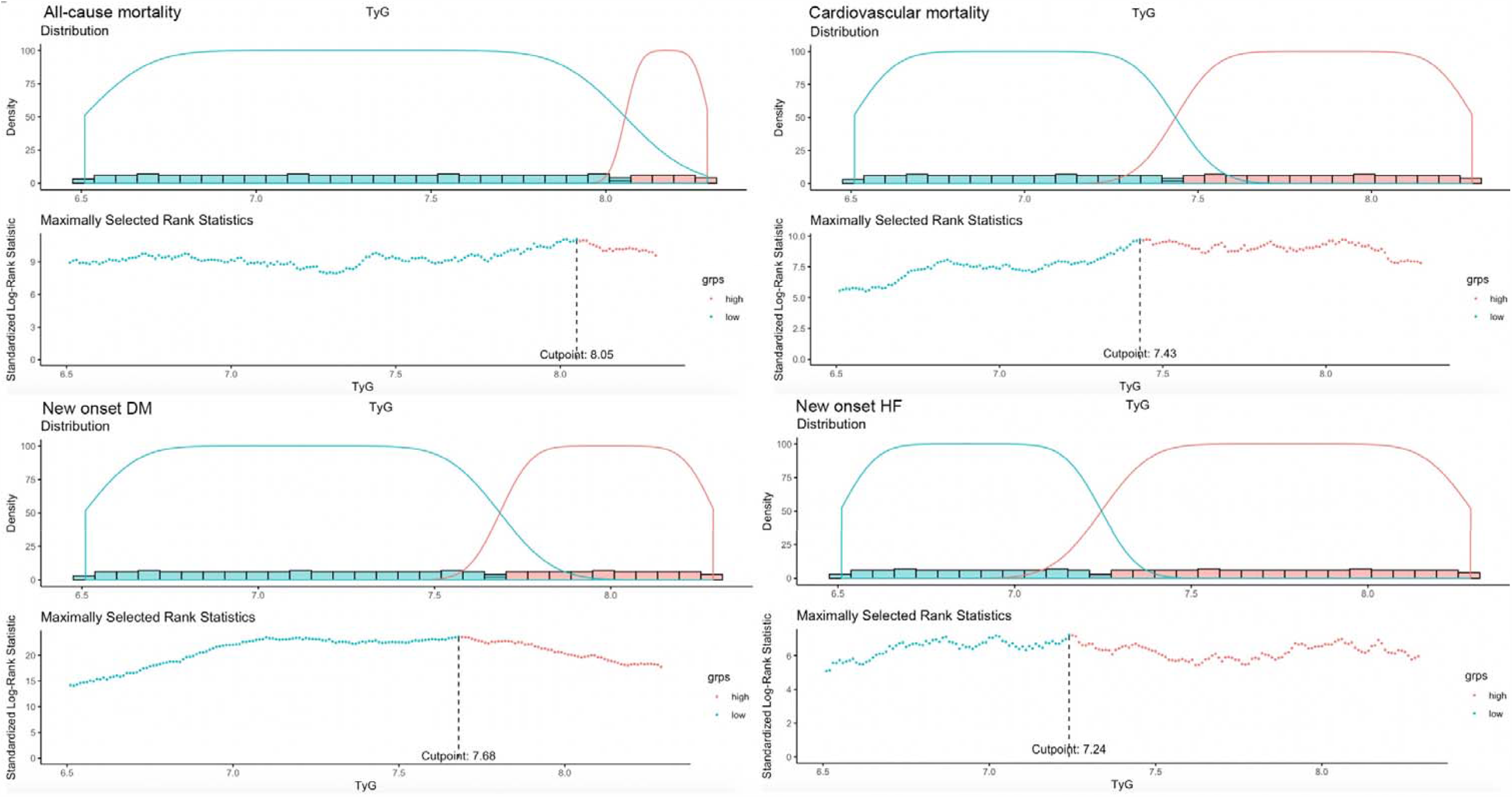
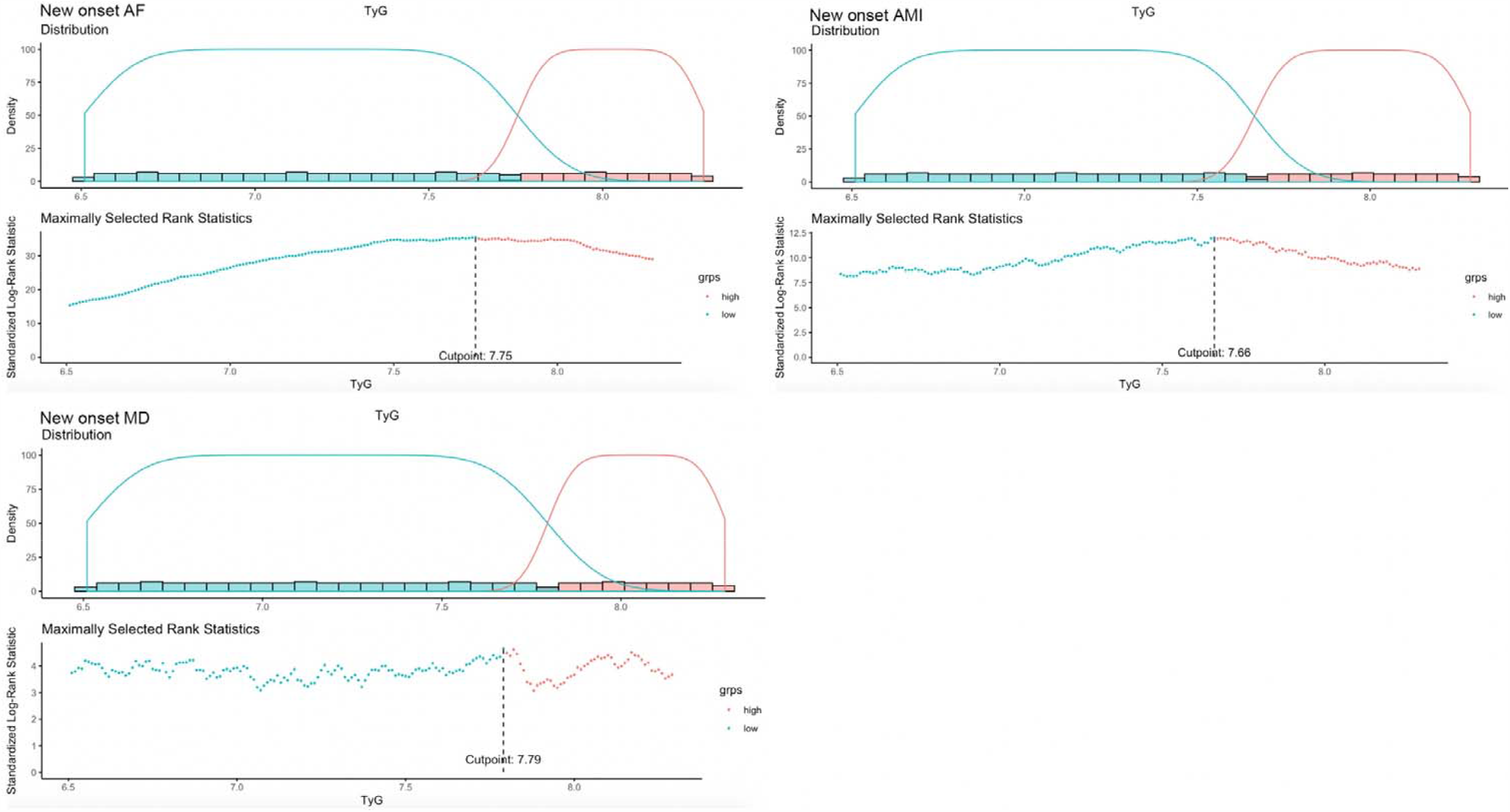
Optimal cutoffs of TyG index to predict primary and secondary outcomes with maximally selected rank statistics approach.

## Results

### Clinical and Biochemical Characteristics

The present cohort consists of 24349 patients (40.2% males, median age of initial triglyceride test: 62.5 years old, IQR: 51.3-71.4, max: 100.97 years old). In total 9103 patients (37.38%) passed away due to all-cause mortality after a median follow-up of 6451 days (IQR: 5142-6911, max: 7517 days), among them 1113 patients (4.57%) died from cardiovascular diseases. Overall 8324 patients (34.18%) developed new onset DM after a median follow up of 6149 days (IQR: 1854-6727, max: 7479 days), 2404 patients (9.87%) developed new onset HF after a median follow up of 6431 days (IQR: 4871-6902, max: 7517 days), 2855 patients (11.72%) developed new onset AF after a median follow up of 6399 days (IQR: 4567-6887, max: 7517 days), 1862 patients (7.64%) developed new onset AMI after a median follow up of 6420 days (IQR: 4808-6895, max: 7517 days), 1275 patients (5.23%) developed new onset VTF after a median follow up of 6445 days (IQR: 5049-6908, max: 7517 days).

The crude incidence rates were 33.11% (N=8062), 33.53% (N=8166) and 33.35% (N=8121) for baseline TyG index in the 1st, 2nd, and 3rd tertiles, respectively, as shown in **Table 1** for the baseline characteristics according to the tertile subgroups of baseline TyG index. There exist significant differences among the three tertiles of TyG index in incidence rates of all-cause mortality (32.69% v.s. 37.68% v.s. 41.74%, P value<0.0001), CAD (3.15% v.s. 4.38% v.s. 6.16%, P value<0.0001), new DM (24.64% v.s. 33.85% v.s. 43.98%, P value<0.0001), new onset HF (7.98% v.s. 10.39% v.s. 11.21%, P value<0.0001), new onset AF (3.80% v.s. 9.57% v.s. 21.74%, P value<0.0001), new onset AMI (5.49% v.s. 7.17% v.s. 10.25%, P value<0.0001) and new onset VTF (4.50% v.s. 5.31% v.s. 5.88%, P value=0.0008).

Significant differences among the three tertile subgroups of patients were also observed in male gender (37.78% v.s. 40.87% v.s. 41.92%, P value=0.0012), baseline age (median: 60.49 years old, IQR: 49.33-70.94 years old; max: 100.97 years old v.s. median: 63.05 years old, IQR: 52.14-71.81; max: 96.23 years old v.s. median: 63.48 years old, IQR: 52.14-71.44; max: 98.94 years old, P value<0.0001), Charlson score (median: 2.0, IQR: 0.0-3.0; max: 11.0 v.s. median: 2.0, IQR: 1.0-3.0; max: 9.0 v.s. median: 2.0, IQR: 1.0-3.0; max: 9.0, P value<0.0001), COPD (0.23% v.s. 0.07% v.s. 0.07%, P value=0.0039), and medication use of ACEI (5.43% v.s. 7.43% v.s. 9.59%, P value<0.0001), calcium channel blockers (9.36% v.s. 11.84% v.s. 12.88%, P value<0.0001), beta blockers (8.71% v.s. 11.36% v.s. 12.17%, P value<0.0001), nitrates (3.88% v.s. 4.65% v.s. 5.27%, P value=0.0003), anti-diabetic drugs (2.70% v.s. 4.83% v.s. 8.81%, P value<0.0001), statins and fibrates (5.80% v.s. 7.46% v.s. 9.77%, P value<0.0001), diuretics for hypertension (4.44% v.s. 6.00% v.s. 6.09%, P value<0.0001), anticoagulants (0.11% v.s. 0.22% v.s. 0.34%, P value=0.0077), antiplatelets (4.70% v.s. 5.55% v.s. 6.44%, P value<0.0001) and lipid-lowering drugs (4.66% v.s. 6.38% v.s. 7.38%, P value<0.0001).

Patients in the 3^rd^ tertile subgroup had significantly lower levels of mean corpuscular volume, mean corpuscular haemoglobin, platelet, reticulocyte, red cell count, hematocrit, urate, albumin, sodium, urea, protein, creatinine, bilirubin and high-density lipoprotein, while had higher levels of lymphocyte, monocyte, neutrophil, white cell count, ALP, aspartate transaminase, ALT, triglyceride, low-density lipoprotein, cholesterol, HbA1c and fast glucose (P value<0.05).

In addition, the baseline characteristics of patients with/without event presentation of new onset DM/AF/HF/AMI/VTF/CAD and all-cause mortality were presented in **Supplementary Tables 2-4**. Patients in the third tertile had a significant larger value of TyG index (median: 6.7, IQR: 6.45-6.88; max: 7.03 v.s. median: 7.31, IQR: 7.17-7.46; max: 7.63 v.s. median: 8.04, IQR: 7.17-8.4; max: 11.61, P value<0.0001). Kaplan-Meier survival curves of the primary and secondary outcomes stratified by deciles of baseline TyG index were presented in **Figure 2**.

### Significant unirivate risk factors of primary and secondary outcomes

Cox-proportional hazard model analysis identified the significant risk factors of primary and secondary outcomes as shown in **Table 2** and **Table 3**. Important predictors of new onset DM include male gender, older patient age, larger Charlson score, comorbidities of liver diseases, IHD, COPD, and PVD (HR>1, P value<0.05); higher levels of mean corpuscular volume, monocyte, neutrophil, white cell count, mean corpuscular haemoglobin, platelet, reticulocyte, hematocrit, potassium, urea, creatinine, alkaline phosphatase, aspartate transaminase, alanine transaminase, bilirubin, triglyceride, HbA1c, fast glucose (HR>1, P value<0.05), and lower levels of lymphocyte, albumin, sodium, protein, low-density lipoprotein, high-density lipoprotein, and cholesterol (HR<1, P value<0.05). Lager TyG index is significantly associated with new onset DM (HR: 1.51, 95% CI: [1.47, 1.55], P vlaue<0.0001). Significances of the stratified decciles and tertiles of TyG index with new onset DM were also presented in **Table 2**. Medication prescriptions of all the mentioned drugs were associated with new onset DM (HR>1, P value<0.05).

Significant risk predictors of new onset HF include male gender, older patient age, larger Charlson score, comorbidities of hypertension, IHD, COPD, PVD, TIA/Ischemic stroke and cancer (HR>1, P value<0.05); higher levels of mean corpuscular volume, monocyte, neutrophil, white cell count, mean corpuscular haemoglobin, platelet, urea, creatinine, alkaline phosphatase, HbA1c, fast glucose (HR>1, P value<0.05), and lower levels of lymphocyte, red cell count, albumin, sodium, protein, alanine transaminase, high-density lipoprotein, and cholesterol (HR<1, P value<0.05). Lager TyG index is significantly associated with new onset HF (HR: 1.27, 95% CI: [1.2, 1.34], P vlaue<0.0001). Significances of the stratified decciles and tertiles of TyG index with new onset DM were also presented. Medication prescriptions of all the mentioned drugs were associated with new onset HF (HR>1, P value<0.05).

Significant predictors of new onset AF included older patient age, larger Chalson score, past comorbidities of hypertension, IHD, COPD, and gastrointestinal bleeding (HR<1, P value<0.05); higher levels of mean corpuscular volume, monocyte, neutrophil, white cell count, mean corpuscular haemoglobin, urea, creatinine, alkaline phosphatase, bilirubin, triglyceride, HbA1c, fast glucose (HR>1, P value<0.05), and lower levels of lymphocyte, platelet, red cell count, potassium, albumin, protein, low-density lipoprotein, and cholesterol (HR<1, P value<0.05). Lager TyG index is significantly associated with new onset AF (HR: 2.36, 95% CI: [2.26, 2.46], P vlaue<0.0001). Significances of the stratified decciles and tertiles of TyG index with new onset AF were also presented. Medication prescriptions of ACEI, ARB, calcium channel blockers, beta blockers, nitrates, antihypertensive drugs, diuretics for hypertension, anticoagulants and antiplatelets were associated with new onset AF (HR>1, P value<0.05).

Significant univariate risk predictors of new onset AMI included male gender, older baseline age, larger Charlson score, past comorbidities of hypertension, IHD, COPD, gastrointestinal diseases, PVD, TIA/Ischemic stroke (HR>1, P value<0.05); higher levels of mean corpuscular volume, monocyte, neutrophil, white cell count, mean corpuscular haemoglobin, platelet, reticulocyte, urea, creatinine, alkaline phosphatase, HbA1c, fast glucose (HR>1, P value<0.05), and lower levels of lymphocyte, platelet, red cell count, sodium, protein and high-density lipoprotein (HR<1, P value<0.05). Lager TyG index is significantly associated with new onset AMI (HR: 1.51, 95% CI: [1.42, 1.6], P vlaue<0.0001). Significances of the stratified decciles and tertiles of TyG index with new onset AMI were also presented. Medication prescriptions of all the mentioned drugs were associated with new onset AMI (HR>1, P value<0.05).

Signifiant risk predictors of new osnet VTF included male gender, older patient age, larger Charlson score, past comorbidities of hypertension, IHD, PVD, TIA/Ischemic stroke (HR>1, P value<0.05); higher levels of mean corpuscular volume, neutrophil, white cell count, mean corpuscular haemoglobin, urea, creatinine, HbA1c, fast glucose (HR>1, P value<0.05), and lower levels of lymphocyte, platelet, red cell count, sodium, protein and high-density lipoprotein (HR<1, P value<0.05). Lager TyG index is significantly associated with new onset VTF (HR: 1.22, 95% CI: [1.13, 1.31], P vlaue<0.0001). Significances of the stratified decciles and tertiles of TyG index with new onset VTF were also presented. Medication prescriptions of all the mentioned drugs were associated with new onset VTF (HR>1, P value<0.05).

### Adjusted associations of baseline TyG index with primary and secondary outcomes

With 1^st^ tertile subgroups of baseline TyG index as a reference, it has been found that the 3^rd^ tertile subgroup were more likely to developed new onset DM/AF/HF/AMI/VTF and meet the outcomes of CAD and all-cause mortality (HR>1, P value<0.05) (**Table 4**), after being adjusted for significant demographics, past comorbidities, and medications in different multivariate Cox analysis models. The optimal cut-off values of baseline TyG index with primary and secondary outcomes were identified with the maximal survival rank statistics approach (**Table 5**). Adjusted multivariate restricted cubic spline models identified the associations between baseline TyG index and HR of primary and secondary outcomes were presented in **Figure 3**.

It has been further confirmed that TyG index above the optimal cut-offs was associated with elevated adverse risk of both the primary and secondary outcomes. Analyses of the associations between baseline TyG index and primary and secondary outcomes with maximal survival rank statistics approach were presented in **Figure 4**. Subgroup characteristics of triglyceride cut-offs with new onset DM and all-cause mortality were presented in **Supplementary Table 2** and **Table 3**, respectively. Basic characteristics of TyG index cut-offs with primary and secondary outcomes were compared in **Supplementary Tables 5-7**.

## Discussion

The major findings of the present study are summarized as follows:

1. Lager TyG index is significantly associated with new onset DM (HR: 1.51, 95% CI: [1.47, 1.55], P vlaue<0.0001), new onset HF (HR: 1.27, 95% CI: [1.2, 1.34], P vlaue<0.0001), new onset AF (HR: 2.36, 95% CI: [2.26, 2.46], P vlaue<0.0001), new onset AMI (HR: 1.51, 95% CI: [1.42, 1.6], P vlaue<0.0001), new onset VTF (HR: 1.22, 95% CI: [1.13, 1.31], P vlaue<0.0001), new onset CAD (HR: 1.56, 95% CI: [1.45, 1.69], P value<0.0001) and all-cause mortality (HR: 1.21, 95% CI: [1.18, 1.25], P vlaue<0.0001);
2. 3^rd^ tertile subgroup (TyG>7.63) were more likely to developed new onset DM/AF/HF/AMI/VTF and meet the outcomes of CAD and all-cause mortality (HR>1, P value<0.05);
3. TyG index and its 3^rd^ tertile remained significant after being adjusted with significant demographics, past comorbidities and medicactions in multivariate cox models (HR>1, P value<0.05);
4. Optimal cut-off values of baseline TyG index were identified and subgroups with TyG index larger than the cut-offs were significantly associated with the primary and secondary outcome (HR>1, P value<0.05);
5. Adjusted multivariate restricted cubic spline models further uncovered the detailed associations between baseline TyG index and HRs of primary and secondary outcomes.

### Limitations

As in other observational studies, this study is limited by potential under-coding of comorbidities, missing data, and coding errors. Additionally, the duration of the complications and the prescribed treatments were not accounted for, which could affect the interpretation of blood pressure value and variability measurements. In addition, this study is conducted based on a Hong Kong cohort, and it is expected that external validity through comparisons with studies from other countries reporting the association between blood pressure variability and adverse outcomes could be conducted for further confirmation. Finally, all patients were older Chinese people, caution should be made when interpreting our findings in younger individuals and other ethnic populations.

## Conclusion

Higher TyG index remained significantly associated with the elevated risk of new onset DM, AF, HF, AMI, VTF, CAD and all-cause mortality after adjustments on demographics, past comorbidities, and medications.

## Data Availability

Data available upon request.

