## Supplementary Appendix for "Triglyceride Glucose Index Predicts Risk of Adverse Cardio-metabolic and Mortality Outcomes Among Chinese Adults: A Territory-Wide Longitudinal Study"

**Supplementary Table 1. ICD9 Codes for comorbidities**

Heart failure	428	428.1	428.2	428.2	428.21	428.22	428.23	428.3	428.3	428.31	428.32	428.33	428.4	428.4	428.41	428.42	428.43	428.9

Congestive heart failure	398.91	402.01	402.11	402.91	404.01	404.03	404.11	404.13	404.91	404.93	428

Atrial fibrillation	427.31	429.4

Liver diseases	456	456.1	456.2	572.2	572.3	572.4	572.8	571.4	571.5	571.6

Endocrine	202.8	200.1	200.12	201.9	204	202.88	200.18	196	204.01	785.6	200.11	200.13	202.85	202.81	196.9	202.82	202	202.87	V10.79	202.84	202.01	196.8	457.1	12.1	196.5	196.2	238.7	196.1	200.14	457.2	V10.61	457	289.3	245.2	457.9	202.93	202.97	757	V10.71	288.8	204.1	202.83	V77.9	237.4	239.7	198.89	623.5	259.9	200.2

Diabetes mellitus	250	250.01	250.02	250.03	250.1	250.11	250.12	250.13	250.2	250.21	250.22	250.23	250.3	250.31	250.32	250.33	250.4	250.41	250.42	250.43	250.5	250.51	250.52	250.53	250.6	250.61	250.62	250.63	250.7	250.71	250.72	250.73	250.8	250.81	250.82	250.83	250.9	250.91	250.92	250.93

Hypertension	401	401.1	401.9	402	402.01	402.1	402.11	402.9	402.91	403	403.01	403.1	403.11	403.9	403.91	404	404.01	404.02	404.03	404.1	404.11	404.12	404.13	404.9	404.91	404.92	404.93	405	405.01	405.09	405.1	405.11	405.19	405.9	405.91	405.99	437.2

AMI	410	410.01	410.02	410.1	410.11	410.12	410.2	410.21	410.22	410.3	410.31	410.32	410.4	410.41	410.42	410.5	410.51	410.52	410.6	410.61	410.62	410.7	410.71	410.72	410.8	410.81	410.82	410.9	410.91	410.92

Stroke/TIA	435	435.1	435.2	435.3	435.8	435.9	433.81	433.91	434	436	437	437.1	433.31	433.01	434.01	434.1	434.11	434.9	434.91	437.2	437.3	437.4	437.5	437.6	437.7	437.8	437.9

IHD	410.01	410.02	410.1	410.11	410.12	410.2	410.21	410.22	410.3	410.31	410.32	410.4	410.41	410.42	410.5	410.51	410.52	410.6	410.61	410.62	410.7	410.71	410.72	410.8	410.81	410.82	410.9	410.91	410.92	411	411.1	411.8	411.81	411.89	413	413.1	413.9	414	414.01	414.02	414.03	414.04	414.05	414.06	414.07	414.1	414.11	414.12	414.19	414.2	414.3	414.4	414.8	414.9	410	412

COPD	490	491	492	493	494	495	496	491.1	491.2	491.21	491.22	491.8	491.9	492.8	493.01	493.02	493.1	493.11	493.12	493.2	493.21	493.22	493.8	493.81	493.82	493.9	493.91	493.92	494.1	495.1	495.2	495.3	495.4	495.5	495.6	495.7	495.8	495.9

Gastrointestinal	153.3	154.1	153.9	569.89	154	153.1	578.9	560.9	569.3	537.89	558.9	562.1	153.6	239	532.3	532.7	535.6	38.42	8.45	153.2	569.49	79.89	532.9	V58.11	569	41.4	152.1	V10.05	787.8	197.4	535.5	V10.06	9	569.83	569.6	153.4	537.3	41.04	569.84	569.81	8.8	535	532	V45.89	V12.72	532.4	V10.09	560.81	235.2	38.49	557.9	569.41	997.4	14.8	787.99	8.46	569.82	537.9	560.1	211.3	556.9	562	536.9	566	V71.9	564.3	V44.4	564.8	562.11	211.2	9.1	8.47	8.5	532.1	535.61	560	565.1	619.1	152.9	568	569.43	152	8.61	596.1	151.4	151.9	151.5	151.8	151.1	456.8	531.7	535.4	531.3	V15.2	211.1	531.9	V10.04	531	531.4	535.1	151.3	230.2	151.6	536.3	535.51	531.1	531.5	537.84	535.01	530.7	535.2	537.6	202.83	558	569.85	153	555.1	562.13	562.12	V76.49	560.2	230.4

PVD	250.7	443.9	443	443.1	443.2	443.21	443.22	443.23	443.24	443.29	443.8	443.81	443.82	443.89	441	443.9	785.4	V43.4

VTF	426	426.12	426.13	426.51	426.52	426.54	427.1	427.4	427.41	427.42	427.5

Gastrointestinal bleeding	531	531.2	531.4	531.6	532	532.2	532.4	532.6	533	533.2	533.4	533.6	534	534.2	534.4	534.6	535.01	535.11	535.21	535.31	535.41	535.51	535.61	535.71	562.02	562.03	562.12	562.13	569.3	569.85	569.86	578	578.1	578.9

Cancer 140-239

**Supplementary Table 2. Basic characteristics of patients with/without new onset DM and patients with/without new onset HF**

* for p≤ 0.05, ** for p ≤ 0.01, *** for p ≤ 0.001

| **Characteristics** | **New onset DM (N=8324) Median (IQR);Max;N or Count(%)** | **No new onset DM (N=16025) Median (IQR);Max;N or Count(%)** | **P value** | **New onset HF (N=2404) Median (IQR);Max;N or Count(%)** | **No new onset HF (N=21945) Median (IQR);Max;N or Count(%)** | **P value** |
| --- | --- | --- | --- | --- | --- | --- |
| TyG index | 7.49(7.05-7.99);11.61;n=8324 | 7.22(6.81-7.7);11.38;n=16025 | <0.0001*** | 7.42(6.99-7.93);10.64;n=2404 | 7.3(6.87-7.8);11.61;n=21945 | <0.0001*** |
| All-cause mortality | 3746(45.00%) | 5357(33.42%) | <0.0001*** | 2008(83.52%) | 7095(32.33%) | <0.0001*** |
| Cardiovascular mortality | 511(6.13%) | 602(3.75%) | <0.0001*** | 335(13.93%) | 778(3.54%) | <0.0001*** |
| New onset DM | 8324(100.00%) | 0(0.00%) | <0.0001*** | 996(41.43%) | 7328(33.39%) | <0.0001*** |
| New onset HF | 996(11.96%) | 1408(8.78%) | <0.0001*** | 2404(100.00%) | 0(0.00%) | <0.0001*** |
| New onset AF | 1047(12.57%) | 1808(11.28%) | 0.0087** | 651(27.07%) | 2204(10.04%) | <0.0001*** |
| New onset AMI | 844(10.13%) | 1018(6.35%) | <0.0001*** | 505(21.00%) | 1357(6.18%) | <0.0001*** |
| New onset ACS | 844(10.13%) | 1018(6.35%) | <0.0001*** | 505(21.00%) | 1357(6.18%) | <0.0001*** |
| New onset VTF | 552(6.63%) | 723(4.51%) | <0.0001*** | 994(41.34%) | 281(1.28%) | <0.0001*** |
| Male gender | 3599(43.23%) | 6190(38.62%) | <0.0001*** | 1061(44.13%) | 8728(39.77%) | 0.0080** |
| Baseline age, year | 64.1(52.8-72.12);100.97;n=8324 | 61.55(50.6-71.02);98.94;n=16025 | <0.0001*** | 70.51(63.1-75.98);96.22;n=2404 | 61.31(50.62-70.53);100.97;n=21945 | <0.0001*** |
| Charlson score | 2.0(1.0-3.0);10.0;n=8324 | 2.0(1.0-3.0);11.0;n=16025 | <0.0001*** | 3.0(2.0-3.0);9.0;n=2404 | 2.0(1.0-3.0);11.0;n=21945 | <0.0001*** |
| Liver diseases | 4(0.04%) | 3(0.01%) | 0.3779 | 0(0.00%) | 7(0.03%) | 0.8088 |
| Endocrine | 13(0.15%) | 18(0.11%) | 0.4718 | 3(0.12%) | 28(0.12%) | 0.7912 |
| Hypertension | 317(3.80%) | 725(4.52%) | 0.0132** | 144(5.99%) | 898(4.09%) | <0.0001*** |
| IHD | 94(1.12%) | 147(0.91%) | 0.1335 | 49(2.03%) | 192(0.87%) | <0.0001*** |
| COPD | 13(0.15%) | 18(0.11%) | 0.4718 | 5(0.20%) | 26(0.11%) | 0.3868 |
| Gastrointestinal | 179(2.15%) | 407(2.53%) | 0.073 | 60(2.49%) | 526(2.39%) | 0.8234 |
| PVD | 8(0.09%) | 5(0.03%) | 0.0741 | 3(0.12%) | 10(0.04%) | 0.2584 |
| TIA/Ischemic stroke | 33(0.39%) | 65(0.40%) | 0.9999 | 16(0.66%) | 82(0.37%) | 0.0494** |
| Gastrointestinal bleeding | 35(0.42%) | 79(0.49%) | 0.4942 | 13(0.54%) | 101(0.46%) | 0.6973 |
| Cancer | 53(0.63%) | 108(0.67%) | 0.7991 | 20(0.83%) | 141(0.64%) | 0.3434 |
| ACEI | 1018(12.22%) | 806(5.02%) | <0.0001*** | 306(12.72%) | 1518(6.91%) | <0.0001*** |
| ARB | 57(0.68%) | 32(0.19%) | <0.0001*** | 19(0.79%) | 70(0.31%) | 0.0006** |
| Calcium channel blockers | 1246(14.96%) | 1522(9.49%) | <0.0001*** | 373(15.51%) | 2395(10.91%) | <0.0001*** |
| Beta blockers | 1064(12.78%) | 1556(9.70%) | <0.0001*** | 307(12.77%) | 2313(10.53%) | 0.0032** |
| Nitrates | 567(6.81%) | 554(3.45%) | <0.0001*** | 202(8.40%) | 919(4.18%) | <0.0001*** |
| Antihypertensive drugs | 482(5.79%) | 574(3.58%) | <0.0001*** | 170(7.07%) | 886(4.03%) | <0.0001*** |
| Anti-Diabetic drugs | 987(11.85%) | 342(2.13%) | <0.0001*** | 193(8.02%) | 1136(5.17%) | <0.0001*** |
| Statins and fibrates | 907(10.89%) | 965(6.02%) | <0.0001*** | 215(8.94%) | 1657(7.55%) | 0.0277** |
| Diuretics for hypertension | 577(6.93%) | 766(4.78%) | <0.0001*** | 175(7.27%) | 1168(5.32%) | 0.0002** |
| Diuretics for heart failure | 209(2.51%) | 148(0.92%) | <0.0001*** | 96(3.99%) | 261(1.18%) | <0.0001*** |
| Anticoagulants | 29(0.34%) | 26(0.16%) | 0.0059** | 9(0.37%) | 46(0.20%) | 0.1662 |
| Antiplatelets | 681(8.18%) | 675(4.21%) | <0.0001*** | 234(9.73%) | 1122(5.11%) | <0.0001*** |
| Lipid-lowering drugs | 717(8.61%) | 780(4.86%) | <0.0001*** | 179(7.44%) | 1318(6.00%) | 0.0103** |
| Mean corpuscular volume, fL | 89.8(86.7-92.7);132.3;n=4368 | 89.6(86.0-92.5);117.0;n=5498 | 0.0014** | 90.1(86.8-93.1);117.0;n=1071 | 89.6(86.3-92.5);132.3;n=8795 | 0.0062** |
| Lymphocyte, x10^9/L | 1.8(1.4-2.3);5.5;n=2782 | 1.8(1.4-2.3);85.28;n=2820 | 0.3411 | 1.71(1.3-2.3);4.9;n=652 | 1.8(1.4-2.3);85.28;n=4950 | 0.0012** |
| Metamyelocyte, x10^9/L | 0.12(0.08-0.29);1.47;n=28 | 0.53(0.18-1.1);2.0;n=17 | 0.0371* | 0.2(0.08-0.43);1.18;n=12 | 0.18(0.11-0.64);2.0;n=33 | 0.8472 |
| Monocyte, x10^9/L | 0.5(0.4-0.64);2.9;n=2780 | 0.5(0.36-0.6);3.22;n=2812 | <0.0001*** | 0.5(0.4-0.7);2.0;n=650 | 0.5(0.4-0.6);3.22;n=4942 | <0.0001*** |
| Neutrophil, x10^9/L | 4.5(3.5-6.4);33.12;n=2778 | 4.1(3.1-5.5);39.48;n=2812 | <0.0001*** | 4.9(3.7-6.6);20.6;n=650 | 4.2(3.3-5.8);39.48;n=4940 | <0.0001*** |
| White blood count, x10^9/L | 7.4(6.1-9.2);36.8;n=4387 | 6.86(5.7-8.37);91.7;n=5537 | <0.0001*** | 7.59(6.18-9.4);24.7;n=1073 | 7.02(5.84-8.6);91.7;n=8851 | <0.0001*** |
| Mean cell haemoglobin, pg | 30.7(29.5-31.8);44.1;n=4368 | 30.5(29.1-31.6);39.6;n=5498 | <0.0001*** | 30.7(29.4-31.9);39.6;n=1071 | 30.5(29.3-31.6);44.1;n=8795 | 0.0044** |
| Myelocyte, x10^9/L | 0.13(0.06-0.4);0.97;n=20 | 0.37(0.24-0.82);2.87;n=10 | 0.0365* | 0.34(0.14-0.45);0.97;n=9 | 0.15(0.1-0.78);2.87;n=21 | 0.6835 |
| Platelet, x10^9/L | 235.0(196.0-279.0);952.0;n=4387 | 238.0(200.0-283.0);1250.0;n=5537 | 0.0009*** | 229.0(191.0-272.0);718.0;n=1073 | 238.0(199.0-283.0);1250.0;n=8851 | <0.0001*** |
| Reticulocyte, x10^9/L | 64.1(44.02-92.7);324.0;n=215 | 50.7(34.1-78.56);194.0;n=153 | 0.0026** | 59.0(43.42-94.55);158.76;n=47 | 58.5(40.6-85.26);324.0;n=321 | 0.4549 |
| Red blood count, x10^12/L | 4.49(4.14-4.82);7.44;n=4366 | 4.4(4.08-4.77);7.62;n=5497 | <0.0001*** | 4.37(4.02-4.74);7.01;n=1070 | 4.44(4.12-4.8);7.62;n=8793 | <0.0001*** |
| Hematocrit, L/L | 0.4(0.37-0.43);0.551;n=600 | 0.39(0.36-0.41);0.541;n=768 | <0.0001*** | 0.39(0.36-0.42);0.49;n=136 | 0.39(0.37-0.42);0.551;n=1232 | 0.7504 |
| K/Potassium, mmol/L | 4.2(3.9-4.6);10.0;n=8216 | 4.2(3.9-4.5);13.3;n=15396 | <0.0001*** | 4.2(3.81-4.55);8.3;n=2366 | 4.2(3.9-4.5);13.3;n=21246 | 0.5009 |
| Albumin, g/L | 42.0(39.2-44.0);58.0;n=5256 | 42.0(40.0-44.0);56.0;n=8111 | 0.1161 | 41.3(39.0-44.0);55.0;n=1365 | 42.0(40.0-44.0);58.0;n=12002 | <0.0001*** |
| Na/Sodium, mmol/L | 140.0(138.6-142.0);168.0;n=8230 | 140.6(139.0-142.0);158.0;n=15440 | <0.0001*** | 140.2(138.8-142.0);168.0;n=2370 | 140.2(139.0-142.0);158.0;n=21300 | 0.6036 |
| Urea, mmol/L | 5.6(4.7-6.8);47.3;n=8232 | 5.5(4.6-6.6);46.4;n=15433 | <0.0001*** | 6.0(4.9-7.4);34.8;n=2368 | 5.5(4.6-6.6);47.3;n=21297 | <0.0001*** |
| Protein, g/L | 75.0(71.0-78.0);93.0;n=5237 | 75.0(71.85-78.0);109.0;n=8068 | 0.6159 | 74.0(70.8-77.7);95.0;n=1362 | 75.0(71.9-78.0);109.0;n=11943 | <0.0001*** |
| Creatinine, umol/L | 85.0(72.0-99.0);621.0;n=8238 | 83.0(71.0-97.0);1509.0;n=15473 | <0.0001*** | 90.0(76.0-106.0);621.0;n=2371 | 83.0(71.0-97.0);1509.0;n=21340 | <0.0001*** |
| Alkaline phosphatase, U/L | 79.0(65.0-96.0);885.0;n=5019 | 76.0(62.0-93.0);827.0;n=7448 | <0.0001*** | 80.0(66.0-97.0);370.0;n=1293 | 77.0(63.0-94.0);885.0;n=11174 | 0.0001*** |
| Aspartate transaminase, U/L | 23.0(18.0-31.0);2640.0;n=1092 | 22.0(18.0-28.0);521.0;n=1348 | 0.0059** | 22.0(18.0-29.0);1516.0;n=249 | 22.0(18.0-28.5);2640.0;n=2191 | 0.7448 |
| Alanine transaminase, U/L | 22.0(15.0-32.0);918.0;n=4188 | 20.0(15.0-29.0);823.0;n=6228 | <0.0001*** | 19.0(14.0-28.0);268.0;n=1122 | 21.0(15.0-30.0);918.0;n=9294 | <0.0001*** |
| Bilirubin, umol/L | 10.0(7.0-13.0);107.0;n=5051 | 9.9(7.2-12.85);216.0;n=7516 | 0.5667 | 9.9(7.0-12.8);48.0;n=1297 | 9.9(7.1-13.0);216.0;n=11270 | 0.5368 |
| Triglyceride, mmol/L | 1.51(1.05-2.19);38.3;n=8324 | 1.35(0.96-1.96);30.3;n=16025 | <0.0001*** | 1.4(1.01-2.03);19.45;n=2404 | 1.4(0.99-2.04);38.3;n=21945 | 0.7092 |
| Low-density lipoprotein, mmol/L | 3.12(2.51-3.79);8.6018;n=7054 | 3.19(2.57-3.83);9.3127;n=13685 | <0.0001*** | 3.12(2.52-3.8);8.0052;n=1967 | 3.17(2.56-3.82);9.3127;n=18772 | 0.1243 |
| High-density lipoprotein, mmol/L | 1.25(1.05-1.5);3.53;n=7276 | 1.32(1.1-1.59);4.14;n=13908 | <0.0001*** | 1.27(1.05-1.55);3.28;n=2011 | 1.3(1.09-1.56);4.14;n=19173 | 0.0005*** |
| Cholesterol, mmol/L | 5.3(4.6-6.0);19.04;n=8308 | 5.33(4.67-6.07);15.88;n=15997 | 0.0005*** | 5.31(4.62-6.04);11.86;n=2400 | 5.31(4.65-6.04);19.04;n=21905 | 0.3512 |
| HbA1c, g/dL | 13.6(12.5-14.5);19.5;n=4218 | 13.2(12.3-13.9);20.0;n=5067 | <0.0001*** | 13.1(12.1-14.2);19.4;n=1006 | 13.3(12.4-14.3);20.0;n=8279 | 0.0007*** |
| Fast glucose, mmol/L | 6.6(5.5-8.57);70.6;n=8324 | 5.5(5.0-6.5);47.1;n=16025 | <0.0001*** | 6.1(5.26-7.8);38.1;n=2404 | 5.7(5.1-7.1);70.6;n=21945 | <0.0001*** |

DM: diabetes mellitus; HF: heart failure; AF: atrial fibrillation; AMI: acute myocardial infarction; VTF: ventricular tachycardia/fibrillation; IHD: ischemic heart disease; COPD: chronic obstructive pulmonary disease; PVD: peripheral vascular disease; TIA: transient ischemic attack; ACEI: angiotensinogen converting enzyme inhibitor; ARB: angiotensin receptor blocker; TyG: triglyceride-glucose

**Supplementary Table 3. Basic characteristics of patients with/without new onset AMI and patients with/without new onset AF**

* for p≤ 0.05, ** for p ≤ 0.01, *** for p ≤ 0.001

| **Characteristics** | **New onset AMI (N=1862) Median (IQR);Max;N or Count(%)** | **No new onset AMI (N=22487) Median (IQR);Max;N or Count(%)** | **P value** | **New onset AF (N=2855) Median (IQR);Max;N or Count(%)** | **No new onset AF (N=21494) Median (IQR);Max;N or Count(%)** | **P value** |
| --- | --- | --- | --- | --- | --- | --- |
| TyG index | 7.53(7.05-8.02);10.47;n=1862 | 7.3(6.87-7.79);11.61;n=22487 | <0.0001*** | 7.84(7.38-8.3);10.93;n=2855 | 7.25(6.84-7.73);11.61;n=21494 | <0.0001*** |
| All-cause mortality | 1317(70.73%) | 7786(34.62%) | <0.0001*** | 1885(66.02%) | 7218(33.58%) | <0.0001*** |
| Cardiovascular mortality | 367(19.70%) | 746(3.31%) | <0.0001*** | 323(11.31%) | 790(3.67%) | <0.0001*** |
| New onset DM | 844(45.32%) | 7480(33.26%) | <0.0001*** | 1047(36.67%) | 7277(33.85%) | 0.0402** |
| New onset HF | 505(27.12%) | 1899(8.44%) | <0.0001*** | 651(22.80%) | 1753(8.15%) | <0.0001*** |
| New onset AF | 461(24.75%) | 2394(10.64%) | <0.0001*** | 2855(100.00%) | 0(0.00%) | <0.0001*** |
| New onset AMI | 1862(100.00%) | 0(0.00%) | <0.0001*** | 461(16.14%) | 1401(6.51%) | <0.0001*** |
| New onset ACS | 1862(100.00%) | 0(0.00%) | <0.0001*** | 461(16.14%) | 1401(6.51%) | <0.0001*** |
| New onset VTF | 318(17.07%) | 957(4.25%) | <0.0001*** | 292(10.22%) | 983(4.57%) | <0.0001*** |
| Male gender | 888(47.69%) | 8901(39.58%) | <0.0001*** | 1125(39.40%) | 8664(40.30%) | 0.5568 |
| Baseline age, year | 69.65(62.56-75.12);100.97;n=1862 | 61.65(50.78-70.91);96.23;n=22487 | <0.0001*** | 70.73(63.68-76.13);95.58;n=2855 | 60.91(50.37-70.32);100.97;n=21494 | <0.0001*** |
| Charlson score | 2.0(2.0-3.0);9.0;n=1862 | 2.0(1.0-3.0);11.0;n=22487 | <0.0001*** | 3.0(2.0-3.0);9.0;n=2855 | 2.0(1.0-3.0);11.0;n=21494 | <0.0001*** |
| Liver diseases | 0(0.00%) | 7(0.03%) | 0.9601 | 0(0.00%) | 7(0.03%) | 0.7064 |
| Endocrine | 1(0.05%) | 30(0.13%) | 0.5567 | 6(0.21%) | 25(0.11%) | 0.2984 |
| Hypertension | 95(5.10%) | 947(4.21%) | 0.0924 | 156(5.46%) | 886(4.12%) | 0.0018** |
| IHD | 40(2.14%) | 201(0.89%) | <0.0001*** | 48(1.68%) | 193(0.89%) | 0.0001** |
| COPD | 4(0.21%) | 27(0.12%) | 0.446 | 6(0.21%) | 25(0.11%) | 0.2984 |
| Gastrointestinal | 26(1.39%) | 560(2.49%) | 0.0047** | 72(2.52%) | 514(2.39%) | 0.7246 |
| PVD | 3(0.16%) | 10(0.04%) | 0.1164 | 2(0.07%) | 11(0.05%) | 0.9831 |
| TIA/Ischemic stroke | 13(0.69%) | 85(0.37%) | 0.058 | 15(0.52%) | 83(0.38%) | 0.3463 |
| Gastrointestinal bleeding | 5(0.26%) | 109(0.48%) | 0.2578 | 23(0.80%) | 91(0.42%) | 0.0081** |
| Cancer | 10(0.53%) | 151(0.67%) | 0.5927 | 19(0.66%) | 142(0.66%) | 0.9259 |
| ACEI | 232(12.45%) | 1592(7.07%) | <0.0001*** | 307(10.75%) | 1517(7.05%) | <0.0001*** |
| ARB | 13(0.69%) | 76(0.33%) | 0.0236** | 21(0.73%) | 68(0.31%) | 0.0010** |
| Calcium channel blockers | 301(16.16%) | 2467(10.97%) | <0.0001*** | 459(16.07%) | 2309(10.74%) | <0.0001*** |
| Beta blockers | 272(14.60%) | 2348(10.44%) | <0.0001*** | 413(14.46%) | 2207(10.26%) | <0.0001*** |
| Nitrates | 186(9.98%) | 935(4.15%) | <0.0001*** | 280(9.80%) | 841(3.91%) | <0.0001*** |
| Antihypertensive drugs | 116(6.22%) | 940(4.18%) | <0.0001*** | 187(6.54%) | 869(4.04%) | <0.0001*** |
| Anti-Diabetic drugs | 157(8.43%) | 1172(5.21%) | <0.0001*** | 151(5.28%) | 1178(5.48%) | 0.7208 |
| Statins and fibrates | 192(10.31%) | 1680(7.47%) | <0.0001*** | 232(8.12%) | 1640(7.63%) | 0.4084 |
| Diuretics for hypertension | 125(6.71%) | 1218(5.41%) | 0.0304** | 215(7.53%) | 1128(5.24%) | <0.0001*** |
| Diuretics for heart failure | 57(3.06%) | 300(1.33%) | <0.0001*** | 103(3.60%) | 254(1.18%) | <0.0001*** |
| Anticoagulants | 7(0.37%) | 48(0.21%) | 0.2455 | 35(1.22%) | 20(0.09%) | <0.0001*** |
| Antiplatelets | 202(10.84%) | 1154(5.13%) | <0.0001*** | 310(10.85%) | 1046(4.86%) | <0.0001*** |
| Lipid-lowering drugs | 141(7.57%) | 1356(6.03%) | 0.0147** | 195(6.83%) | 1302(6.05%) | 0.1408 |
| Mean corpuscular volume, fL | 90.1(86.95-93.0);117.0;n=843 | 89.6(86.3-92.6);132.3;n=9023 | 0.013* | 90.3(86.7-93.5);117.0;n=1237 | 89.6(86.3-92.5);132.3;n=8629 | <0.0001*** |
| Lymphocyte, x10^9/L | 1.8(1.4-2.3);5.5;n=495 | 1.8(1.4-2.3);85.28;n=5107 | 0.5134 | 1.8(1.4-2.3);5.5;n=737 | 1.8(1.4-2.3);85.28;n=4865 | 0.0226 |
| Metamyelocyte, x10^9/L | 0.14(0.08-0.48);0.76;n=4 | 0.19(0.11-0.62);2.0;n=41 | 0.6898 | 0.11(0.08-0.19);1.47;n=9 | 0.2(0.12-0.62);2.0;n=36 | 0.2013 |
| Monocyte, x10^9/L | 0.5(0.4-0.7);3.18;n=494 | 0.5(0.4-0.6);3.22;n=5098 | <0.0001*** | 0.5(0.4-0.7);2.0;n=736 | 0.5(0.4-0.6);3.22;n=4856 | <0.0001*** |
| Neutrophil, x10^9/L | 4.8(3.7-6.7);26.7;n=494 | 4.2(3.3-5.82);39.48;n=5096 | <0.0001*** | 4.6(3.5-6.5);33.12;n=735 | 4.2(3.3-5.8);39.48;n=4855 | <0.0001*** |
| White blood count, x10^9/L | 7.7(6.4-9.52);31.2;n=850 | 7.02(5.8-8.6);91.7;n=9074 | <0.0001*** | 7.31(6.0-9.0);36.8;n=1244 | 7.08(5.88-8.68);91.7;n=8680 | 0.0004*** |
| Mean cell haemoglobin, pg | 30.7(29.5-31.8);39.6;n=843 | 30.5(29.3-31.7);44.1;n=9023 | 0.009** | 30.7(29.4-32.0);39.6;n=1237 | 30.5(29.3-31.6);44.1;n=8629 | 0.0001*** |
| Myelocyte, x10^9/L | 0.35(0.35-0.35);0.4;n=2 | 0.18(0.1-0.69);2.87;n=28 | 0.5886 | 0.2(0.04-0.24);0.34;n=5 | 0.3(0.11-0.78);2.87;n=25 | 0.2102 |
| Platelet, x10^9/L | 229.5(196.0-273.0);505.0;n=850 | 238.0(198.0-282.0);1250.0;n=9074 | 0.0132 | 222.0(183.0-264.0);935.0;n=1244 | 239.0(200.0-283.0);1250.0;n=8680 | <0.0001*** |
| Reticulocyte, x10^9/L | 80.4(47.6-97.56);307.58;n=33 | 57.2(40.46-85.02);324.0;n=335 | 0.0511 | 71.04(43.0-92.38);192.0;n=55 | 56.0(40.6-84.74);324.0;n=313 | 0.1068 |
| Red blood count, x10^12/L | 4.4(4.0-4.79);7.01;n=843 | 4.44(4.11-4.8);7.62;n=9020 | 0.0138 | 4.37(4.03-4.77);7.44;n=1237 | 4.44(4.12-4.8);7.62;n=8626 | 0.0003*** |
| Hematocrit, L/L | 0.39(0.36-0.42);0.551;n=126 | 0.39(0.37-0.42);0.541;n=1242 | 0.3142 | 0.39(0.36-0.42);0.551;n=172 | 0.4(0.37-0.42);0.541;n=1196 | 0.0695 |
| K/Potassium, mmol/L | 4.2(3.86-4.53);6.6;n=1832 | 4.2(3.9-4.5);13.3;n=21780 | 0.6108 | 4.2(3.8-4.5);7.9;n=2804 | 4.2(3.9-4.5);13.3;n=20808 | <0.0001*** |
| Albumin, g/L | 41.4(39.0-44.0);53.0;n=1049 | 42.0(40.0-44.0);58.0;n=12318 | <0.0001*** | 41.0(39.0-44.0);54.0;n=1580 | 42.0(40.0-44.0);58.0;n=11787 | <0.0001*** |
| Na/Sodium, mmol/L | 140.0(138.4-142.0);152.0;n=1833 | 140.3(139.0-142.0);168.0;n=21837 | 0.0309* | 141.0(139.0-142.0);155.0;n=2810 | 140.0(139.0-142.0);168.0;n=20860 | 0.0003*** |
| Urea, mmol/L | 6.0(5.0-7.45);46.4;n=1835 | 5.5(4.6-6.6);47.3;n=21830 | <0.0001*** | 5.9(4.9-7.2);46.4;n=2810 | 5.5(4.6-6.6);47.3;n=20855 | <0.0001*** |
| Protein, g/L | 74.0(71.0-78.0);95.0;n=1045 | 75.0(71.6-78.0);109.0;n=12260 | 0.0019** | 74.0(71.0-78.0);95.0;n=1572 | 75.0(72.0-78.0);109.0;n=11733 | <0.0001*** |
| Creatinine, umol/L | 91.0(78.0-107.0);1509.0;n=1839 | 83.0(71.0-97.0);1476.0;n=21872 | <0.0001*** | 87.0(75.0-103.0);1509.0;n=2814 | 83.0(71.0-97.0);1476.0;n=20897 | <0.0001*** |
| Alkaline phosphatase, U/L | 81.0(67.0-98.0);428.0;n=987 | 77.0(63.0-94.0);885.0;n=11480 | <0.0001*** | 80.0(66.0-98.0);885.0;n=1476 | 77.0(63.0-94.0);827.0;n=10991 | <0.0001*** |
| Aspartate transaminase, U/L | 22.0(18.0-30.0);1516.0;n=217 | 22.0(18.0-28.0);2640.0;n=2223 | 0.8496 | 23.0(18.0-29.5);922.0;n=322 | 22.0(18.0-28.0);2640.0;n=2118 | 0.3114 |
| Alanine transaminase, U/L | 20.0(15.0-30.0);268.0;n=833 | 21.0(15.0-30.0);918.0;n=9583 | 0.4726 | 20.0(14.0-28.0);456.0;n=1272 | 21.0(15.0-30.0);918.0;n=9144 | 0.001** |
| Bilirubin, umol/L | 10.0(7.0-13.0);59.0;n=994 | 9.9(7.1-13.0);216.0;n=11573 | 0.4463 | 10.0(7.9-13.7);216.0;n=1483 | 9.75(7.0-12.8);140.0;n=11084 | <0.0001*** |
| Triglyceride, mmol/L | 1.49(1.05-2.17);18.74;n=1862 | 1.39(0.98-2.03);38.3;n=22487 | <0.0001*** | 1.35(0.96-1.92);21.89;n=2855 | 1.4(1.0-2.05);38.3;n=21494 | <0.0001*** |
| Low-density lipoprotein, mmol/L | 3.2(2.55-3.89);8.0052;n=1603 | 3.17(2.55-3.81);9.3127;n=19136 | 0.2242 | 3.09(2.46-3.76);7.7791;n=2356 | 3.18(2.56-3.82);9.3127;n=18383 | <0.0001*** |
| High-density lipoprotein, mmol/L | 1.22(1.01-1.47);3.05;n=1642 | 1.3(1.09-1.57);4.14;n=19542 | <0.0001*** | 1.31(1.1-1.57);3.1;n=2398 | 1.3(1.08-1.56);4.14;n=18786 | 0.3578 |
| Cholesterol, mmol/L | 5.37(4.69-6.1);10.7;n=1861 | 5.3(4.64-6.04);19.04;n=22444 | 0.0568 | 5.22(4.56-5.98);10.63;n=2846 | 5.32(4.66-6.06);19.04;n=21459 | <0.0001*** |
| HbA1c, g/dL | 13.4(12.2-14.3);19.4;n=805 | 13.3(12.4-14.3);20.0;n=8480 | 0.7829 | 13.2(12.2-14.2);20.0;n=1163 | 13.3(12.4-14.3);19.5;n=8122 | 0.0186 |
| Fast glucose, mmol/L | 6.3(5.3-8.45);35.3;n=1862 | 5.7(5.1-7.1);70.6;n=22487 | <0.0001*** | 5.9(5.29-7.3);65.6;n=2855 | 5.7(5.1-7.2);70.6;n=21494 | <0.0001*** |

DM: diabetes mellitus; HF: heart failure; AF: atrial fibrillation; AMI: acute myocardial infarction; VTF: ventricular tachycardia/fibrillation; IHD: ischemic heart disease; COPD: chronic obstructive pulmonary disease; PVD: peripheral vascular disease; TIA: transient ischemic attack; ACEI: angiotensinogen converting enzyme inhibitor; ARB: angiotensin receptor blocker; TyG: triglyceride-glucose

**Supplementary Table 4. Basic characteristics of patients with/without new onset VTF**

* for p≤ 0.05, ** for p ≤ 0.01, *** for p ≤ 0.001

| **Characteristics** | **New onset VTF (N=1275) Median (IQR);Max;N or Count(%)** | **No new onset VTF (N=23074) Median (IQR);Max;N or Count(%)** | **P value** |
| --- | --- | --- | --- |
| TyG index | 7.4(6.96-7.88);10.5;n=1275 | 7.31(6.88-7.8);11.61;n=23074 | <0.0001*** |
| All-cause mortality | 1138(89.25%) | 7965(34.51%) | <0.0001*** |
| Cardiovascular mortality | 246(19.29%) | 867(3.75%) | <0.0001*** |
| New onset DM | 552(43.29%) | 7772(33.68%) | <0.0001*** |
| New onset HF | 994(77.96%) | 1410(6.11%) | <0.0001*** |
| New onset AF | 292(22.90%) | 2563(11.10%) | <0.0001*** |
| New onset AMI | 318(24.94%) | 1544(6.69%) | <0.0001*** |
| New onset ACS | 318(24.94%) | 1544(6.69%) | <0.0001*** |
| New onset VTF | 1275(100.00%) | 0(0.00%) | <0.0001*** |
| Male gender | 659(51.68%) | 9130(39.56%) | <0.0001*** |
| Baseline age, year | 70.89(64.39-75.93);96.22;n=1275 | 61.82(50.89-70.97);100.97;n=23074 | <0.0001*** |
| Charlson score | 3.0(2.0-3.0);5.0;n=1275 | 2.0(1.0-3.0);11.0;n=23074 | <0.0001*** |
| Liver diseases | 0(0.00%) | 7(0.03%) | 0.8207 |
| Endocrine | 1(0.07%) | 30(0.13%) | 0.9213 |
| Hypertension | 74(5.80%) | 968(4.19%) | 0.0105** |
| IHD | 18(1.41%) | 223(0.96%) | 0.1615 |
| COPD | 0(0.00%) | 31(0.13%) | 0.3654 |
| Gastrointestinal | 32(2.50%) | 554(2.40%) | 0.8831 |
| PVD | 3(0.23%) | 10(0.04%) | 0.0237** |
| TIA/Ischemic stroke | 11(0.86%) | 87(0.37%) | 0.0154** |
| Gastrointestinal bleeding | 8(0.62%) | 106(0.45%) | 0.5218 |
| Cancer | 4(0.31%) | 157(0.68%) | 0.1654 |
| ACEI | 173(13.56%) | 1651(7.15%) | <0.0001*** |
| ARB | 9(0.70%) | 80(0.34%) | 0.0688 |
| Calcium channel blockers | 193(15.13%) | 2575(11.15%) | 0.0002** |
| Beta blockers | 162(12.70%) | 2458(10.65%) | 0.0450** |
| Nitrates | 97(7.60%) | 1024(4.43%) | <0.0001*** |
| Antihypertensive drugs | 95(7.45%) | 961(4.16%) | <0.0001*** |
| Anti-Diabetic drugs | 99(7.76%) | 1230(5.33%) | 0.0006** |
| Statins and fibrates | 121(9.49%) | 1751(7.58%) | 0.0260** |
| Diuretics for hypertension | 85(6.66%) | 1258(5.45%) | 0.0932 |
| Diuretics for heart failure | 37(2.90%) | 320(1.38%) | <0.0001*** |
| Anticoagulants | 6(0.47%) | 49(0.21%) | 0.1137 |
| Antiplatelets | 125(9.80%) | 1231(5.33%) | <0.0001*** |
| Lipid-lowering drugs | 100(7.84%) | 1397(6.05%) | 0.0185** |
| Mean corpuscular volume, fL | 90.35(87.15-93.2);117.0;n=568 | 89.6(86.3-92.6);132.3;n=9298 | 0.0002*** |
| Lymphocyte, x10^9/L | 1.78(1.3-2.3);4.5;n=338 | 1.8(1.4-2.3);85.28;n=5264 | 0.0523 |
| Metamyelocyte, x10^9/L | 0.16(0.1-0.24);0.3;n=6 | 0.19(0.1-0.64);2.0;n=39 | 0.4226 |
| Monocyte, x10^9/L | 0.5(0.4-0.6);2.0;n=337 | 0.5(0.4-0.6);3.22;n=5255 | 0.3741 |
| Neutrophil, x10^9/L | 4.7(3.6-6.2);20.3;n=337 | 4.2(3.3-5.9);39.48;n=5253 | 0.0022** |
| White blood count, x10^9/L | 7.3(6.01-9.1);20.0;n=570 | 7.1(5.9-8.7);91.7;n=9354 | 0.0168* |
| Mean cell haemoglobin, pg | 30.8(29.6-32.0);39.6;n=568 | 30.5(29.3-31.7);44.1;n=9298 | 0.001*8 |
| Myelocyte, x10^9/L | 0.37(0.18-0.43);0.45;n=4 | 0.18(0.11-0.69);2.87;n=26 | 0.9513 |
| Platelet, x10^9/L | 226.0(186.5-266.0);511.0;n=570 | 238.0(199.0-282.0);1250.0;n=9354 | <0.0001*** |
| Reticulocyte, x10^9/L | 59.95(42.98-94.55);158.76;n=28 | 58.65(40.46-85.38);324.0;n=340 | 0.437 |
| Red blood count, x10^12/L | 4.37(4.01-4.79);6.71;n=568 | 4.44(4.11-4.8);7.62;n=9295 | 0.0217* |
| Hematocrit, L/L | 0.38(0.36-0.41);0.48;n=81 | 0.39(0.37-0.42);0.551;n=1287 | 0.1809 |
| K/Potassium, mmol/L | 4.2(3.9-4.56);6.8;n=1252 | 4.2(3.9-4.5);13.3;n=22360 | 0.3134 |
| Albumin, g/L | 41.6(39.0-44.0);55.0;n=717 | 42.0(39.9-44.0);58.0;n=12650 | 0.0191* |
| Na/Sodium, mmol/L | 140.55(139.0-142.0);152.0;n=1254 | 140.2(139.0-142.0);168.0;n=22416 | 0.9646 |
| Urea, mmol/L | 6.1(5.0-7.5);31.6;n=1253 | 5.5(4.6-6.6);47.3;n=22412 | <0.0001*** |
| Protein, g/L | 74.0(71.0-77.5);89.0;n=714 | 75.0(71.6-78.0);109.0;n=12591 | 0.001** |
| Creatinine, umol/L | 91.0(78.0-107.0);434.0;n=1256 | 83.0(71.0-97.0);1509.0;n=22455 | <0.0001*** |
| Alkaline phosphatase, U/L | 81.0(67.0-97.0);300.0;n=678 | 77.0(63.0-94.0);885.0;n=11789 | 0.0004*** |
| Aspartate transaminase, U/L | 21.0(18.0-26.0);1516.0;n=130 | 22.0(18.0-29.0);2640.0;n=2310 | 0.0686 |
| Alanine transaminase, U/L | 19.0(14.0-27.5);215.0;n=572 | 21.0(15.0-30.0);918.0;n=9844 | 0.0002*** |
| Bilirubin, umol/L | 9.9(7.6-12.2);38.0;n=681 | 9.9(7.0-13.0);216.0;n=11886 | 0.7519 |
| Triglyceride, mmol/L | 1.4(1.0-2.01);11.19;n=1275 | 1.4(0.99-2.04);38.3;n=23074 | 0.9555 |
| Low-density lipoprotein, mmol/L | 3.15(2.58-3.85);8.0052;n=1046 | 3.17(2.55-3.82);9.3;n=19693 | 0.9018 |
| High-density lipoprotein, mmol/L | 1.25(1.05-1.52);3.28;n=1072 | 1.3(1.09-1.56);4.14;n=20112 | 0.0002*** |
| Cholesterol, mmol/L | 5.3(4.63-6.02);10.77;n=1273 | 5.31(4.64-6.05);19.04;n=23032 | 0.3109 |
| HbA1c, g/dL | 13.3(12.2-14.4);19.4;n=535 | 13.3(12.4-14.2);20.0;n=8750 | 0.8447 |
| Fast glucose, mmol/L | 6.1(5.29-7.8);38.4;n=1275 | 5.7(5.1-7.2);70.6;n=23074 | <0.0001*** |

DM: diabetes mellitus; HF: heart failure; AF: atrial fibrillation; AMI: acute myocardial infarction; VTF: ventricular tachycardia/fibrillation; IHD: ischemic heart disease; COPD: chronic obstructive pulmonary disease; PVD: peripheral vascular disease; TIA: transient ischemic attack; ACEI: angiotensinogen converting enzyme inhibitor; ARB: angiotensin receptor blocker; TyG: triglyceride-glucose

**Supplementary Table 4. Basic characteristics of TyG index cut-off with all-cause mortality and CAD**

* for p≤ 0.05, ** for p ≤ 0.01, *** for p ≤ 0.001

| **Characteristics** | **TyG>8.05 (N=3940) Median (IQR);Max;N or Count(%)** | **TyG**$\boldsymbol{\leq}$**8.05 (N=20409) Median (IQR);Max;N or Count(%)** | **P value** | **TyG>7.43 (N=10506) Median (IQR);Max;N or Count(%)** | **TyG**$\boldsymbol{\leq}$**7.43 (N=13843) Median (IQR);Max;N or Count(%)** | **P value** |
| --- | --- | --- | --- | --- | --- | --- |
| Male gender | 1697(43.07%) | 8092(39.64%) | 0.0098** | 4387(41.75%) | 5402(39.02%) | 0.0051** |
| Baseline age, year | 64.07(52.96-71.54);98.94;n=3940 | 62.17(50.98-71.34);100.97;n=20409 | <0.0001*** | 63.54(52.73-71.51);98.94;n=10506 | 61.54(50.36-71.29);100.97;n=13843 | <0.0001*** |
| Charlson score | 2.0(1.0-3.0);6.0;n=3940 | 2.0(1.0-3.0);11.0;n=20409 | <0.0001*** | 2.0(1.0-3.0);9.0;n=10506 | 2.0(1.0-3.0);11.0;n=13843 | <0.0001*** |
| Liver diseases | 1(0.02%) | 6(0.02%) | 0.7061 | 4(0.03%) | 3(0.02%) | 0.7145 |
| Endocrine | 4(0.10%) | 27(0.13%) | 0.8016 | 13(0.12%) | 18(0.13%) | 0.9639 |
| Hypertension | 158(4.01%) | 884(4.33%) | 0.4055 | 433(4.12%) | 609(4.39%) | 0.3247 |
| IHD | 41(1.04%) | 200(0.97%) | 0.7943 | 103(0.98%) | 138(0.99%) | 0.9504 |
| COPD | 4(0.10%) | 27(0.13%) | 0.8016 | 9(0.08%) | 22(0.15%) | 0.1602 |
| Gastrointestinal | 86(2.18%) | 500(2.44%) | 0.3565 | 237(2.25%) | 349(2.52%) | 0.2062 |
| PVD | 1(0.02%) | 12(0.05%) | 0.6496 | 4(0.03%) | 9(0.06%) | 0.5347 |
| TIA/Ischemic stroke | 15(0.38%) | 83(0.40%) | 0.9224 | 46(0.43%) | 52(0.37%) | 0.5131 |
| Gastrointestinal bleeding | 16(0.40%) | 98(0.48%) | 0.6217 | 54(0.51%) | 60(0.43%) | 0.4162 |
| Cancer | 27(0.68%) | 134(0.65%) | 0.9244 | 64(0.60%) | 97(0.70%) | 0.4311 |
| ACEI | 414(10.50%) | 1410(6.90%) | <0.0001*** | 985(9.37%) | 839(6.06%) | <0.0001*** |
| ARB | 19(0.48%) | 70(0.34%) | 0.2394 | 43(0.40%) | 46(0.33%) | 0.3815 |
| Calcium channel blockers | 521(13.22%) | 2247(11.00%) | 0.0004*** | 1349(12.84%) | 1419(10.25%) | <0.0001*** |
| Beta blockers | 506(12.84%) | 2114(10.35%) | <0.0001*** | 1273(12.11%) | 1347(9.73%) | <0.0001*** |
| Nitrates | 231(5.86%) | 890(4.36%) | 0.0001*** | 554(5.27%) | 567(4.09%) | <0.0001*** |
| Antihypertensive drugs | 184(4.67%) | 872(4.27%) | 0.3032 | 461(4.38%) | 595(4.29%) | 0.7685 |
| Anti-Diabetic drugs | 412(10.45%) | 917(4.49%) | <0.0001*** | 849(8.08%) | 480(3.46%) | <0.0001*** |
| Statins and fibrates | 436(11.06%) | 1436(7.03%) | <0.0001*** | 994(9.46%) | 878(6.34%) | <0.0001*** |
| Diuretics for hypertension | 264(6.70%) | 1079(5.28%) | 0.0009*** | 651(6.19%) | 692(4.99%) | 0.0001*** |
| Diuretics for heart failure | 73(1.85%) | 284(1.39%) | 0.0359* | 174(1.65%) | 183(1.32%) | 0.0391* |
| Anticoagulants | 18(0.45%) | 37(0.18%) | 0.0017** | 35(0.33%) | 20(0.14%) | 0.0034** |
| Antiplatelets | 268(6.80%) | 1088(5.33%) | 0.0006*** | 676(6.43%) | 680(4.91%) | <0.0001*** |
| Lipid-lowering drugs | 311(7.89%) | 1186(5.81%) | <0.0001*** | 762(7.25%) | 735(5.30%) | <0.0001*** |
| Mean corpuscular volume, fL | 89.0(85.4-92.2);108.0;n=1664 | 89.8(86.5-92.7);132.3;n=8202 | <0.0001*** | 89.1(85.8-92.2);132.3;n=4315 | 90.1(86.8-92.9);119.5;n=5551 | <0.0001*** |
| Lymphocyte, x10^9/L | 2.0(1.5-2.5);5.2;n=947 | 1.8(1.4-2.3);85.28;n=4655 | <0.0001*** | 1.9(1.5-2.46);5.5;n=2442 | 1.8(1.3-2.2);85.28;n=3160 | <0.0001*** |
| Metamyelocyte, x10^9/L | 0.2(0.12-0.59);2.0;n=15 | 0.17(0.08-0.58);1.79;n=30 | 0.3173 | 0.2(0.12-0.62);2.0;n=30 | 0.12(0.06-0.4);1.47;n=15 | 0.2474 |
| Monocyte, x10^9/L | 0.5(0.4-0.64);3.22;n=944 | 0.5(0.4-0.6);3.18;n=4648 | <0.0001*** | 0.5(0.4-0.63);3.22;n=2437 | 0.5(0.39-0.6);3.18;n=3155 | <0.0001*** |
| Neutrophil, x10^9/L | 4.6(3.6-6.5);28.2;n=944 | 4.2(3.2-5.8);39.48;n=4646 | <0.0001*** | 4.5(3.5-6.3);39.48;n=2437 | 4.1(3.12-5.63);36.67;n=3153 | <0.0001*** |
| White blood count, x10^9/L | 7.7(6.4-9.5);30.4;n=1673 | 7.0(5.8-8.5);91.7;n=8251 | <0.0001*** | 7.54(6.27-9.25);47.0;n=4345 | 6.8(5.64-8.3);91.7;n=5579 | <0.0001*** |
| Mean cell haemoglobin, pg | 30.5(29.15-31.7);38.0;n=1664 | 30.6(29.3-31.7);44.1;n=8202 | 0.6791 | 30.5(29.2-31.6);44.1;n=4315 | 30.6(29.4-31.7);41.5;n=5551 | <0.0001*** |
| Myelocyte, x10^9/L | 0.37(0.16-0.8);0.97;n=8 | 0.15(0.1-0.4);2.87;n=22 | 0.2804 | 0.34(0.13-0.83);2.87;n=17 | 0.12(0.1-0.34);0.81;n=13 | 0.1941 |
| Platelet, x10^9/L | 241.0(203.0-284.0);749.0;n=1673 | 236.0(197.0-281.0);1250.0;n=8251 | 0.0175* | 241.0(203.0-284.0);1250.0;n=4345 | 233.0(195.0-279.0);957.0;n=5579 | <0.0001*** |
| Reticulocyte, x10^9/L | 73.44(49.8-103.0);307.58;n=73 | 54.1(38.7-81.3);324.0;n=295 | 0.0005*** | 69.9(43.94-92.7);307.58;n=163 | 52.3(36.8-78.88);324.0;n=205 | 0.0003*** |
| Red blood count, x10^12/L | 4.51(4.15-4.93);7.62;n=1664 | 4.42(4.1-4.77);7.61;n=8199 | <0.0001*** | 4.49(4.15-4.86);7.62;n=4315 | 4.39(4.08-4.74);7.61;n=5548 | <0.0001*** |
| Hematocrit, L/L | 0.4(0.37-0.43);0.551;n=225 | 0.39(0.37-0.42);0.519;n=1143 | 0.0096** | 0.4(0.37-0.42);0.551;n=590 | 0.39(0.37-0.42);0.519;n=778 | 0.0473* |
| K/Potassium, mmol/L | 4.2(3.84-4.51);6.6;n=3879 | 4.2(3.9-4.5);13.3;n=19733 | 0.666 | 4.2(3.87-4.5);8.1;n=10291 | 4.2(3.9-4.5);13.3;n=13321 | 0.2815 |
| Albumin, g/L | 42.0(39.9-44.0);56.0;n=2201 | 42.0(39.9-44.0);58.0;n=11166 | 0.3613 | 42.0(40.0-44.0);56.0;n=5826 | 42.0(39.8-44.0);58.0;n=7541 | <0.0001*** |
| Na/Sodium, mmol/L | 140.0(138.0-142.0);168.0;n=3886 | 141.0(139.0-142.0);158.0;n=19784 | <0.0001*** | 140.0(138.0-142.0);168.0;n=10313 | 141.0(139.0-142.0);152.0;n=13357 | <0.0001*** |
| Urea, mmol/L | 5.6(4.7-6.9);46.4;n=3886 | 5.5(4.6-6.6);47.3;n=19779 | <0.0001*** | 5.6(4.6-6.8);46.4;n=10315 | 5.5(4.6-6.6);47.3;n=13350 | <0.0001*** |
| Protein, g/L | 75.0(72.0-79.0);109.0;n=2193 | 75.0(71.0-78.0);109.0;n=11112 | <0.0001*** | 75.0(72.0-79.0);109.0;n=5804 | 74.0(71.0-77.8);100.0;n=7501 | <0.0001*** |
| Creatinine, umol/L | 86.0(73.0-101.0);1509.0;n=3892 | 83.0(71.0-97.0);1476.0;n=19819 | <0.0001*** | 85.0(73.0-99.0);1509.0;n=10329 | 82.0(71.0-97.0);629.0;n=13382 | <0.0001*** |
| Alkaline phosphatase, U/L | 82.0(67.0-100.0);885.0;n=2050 | 76.0(63.0-93.0);827.0;n=10417 | <0.0001*** | 80.0(66.0-97.0);885.0;n=5440 | 75.0(62.0-92.0);827.0;n=7027 | <0.0001*** |
| Aspartate transaminase, U/L | 23.0(18.0-30.0);2640.0;n=423 | 22.0(18.0-28.0);1787.0;n=2017 | 0.2188 | 23.0(18.0-30.0);2640.0;n=1083 | 22.0(18.0-28.0);1787.0;n=1357 | 0.0335* |
| Alanine transaminase, U/L | 24.0(17.0-34.0);823.0;n=1657 | 20.0(15.0-29.0);918.0;n=8759 | <0.0001*** | 23.0(16.0-33.0);918.0;n=4456 | 19.0(14.0-27.0);700.0;n=5960 | <0.0001*** |
| Bilirubin, umol/L | 9.0(7.0-12.0);70.0;n=2065 | 10.0(7.3-13.0);216.0;n=10502 | <0.0001*** | 9.4(7.0-12.6);107.0;n=5482 | 10.0(7.4-13.0);216.0;n=7085 | <0.0001*** |
| Triglyceride, mmol/L | 2.84(1.98-4.04);38.3;n=3940 | 1.28(0.93-1.75);6.2;n=20409 | <0.0001*** | 2.16(1.64-2.86);38.3;n=10506 | 1.09(0.83-1.39);3.0;n=13843 | <0.0001*** |
| Low-density lipoprotein, mmol/L | 3.08(2.42-3.76);7.7791;n=3187 | 3.19(2.58-3.83);9.3127;n=17552 | <0.0001*** | 3.19(2.55-3.85);9.3045;n=8965 | 3.15(2.56-3.79);9.3127;n=11774 | 0.0655 |
| High-density lipoprotein, mmol/L | 1.11(0.95-1.32);3.38;n=3551 | 1.34(1.12-1.6);4.14;n=17633 | <0.0001*** | 1.17(1.0-1.38);3.38;n=9366 | 1.41(1.19-1.68);4.14;n=11818 | <0.0001*** |
| Cholesterol, mmol/L | 5.6(4.9-6.39);19.04;n=3931 | 5.27(4.6-5.98);12.205;n=20374 | <0.0001*** | 5.51(4.82-6.25);19.04;n=10486 | 5.17(4.52-5.87);11.5;n=13819 | <0.0001*** |
| HbA1c, g/dL | 13.5(12.5-14.6);18.4;n=1564 | 13.3(12.3-14.2);20.0;n=7721 | <0.0001*** | 13.4(12.4-14.4);20.0;n=4064 | 13.2(12.3-14.1);19.5;n=5221 | <0.0001*** |
| Fast glucose, mmol/L | 8.3(6.1-12.0);70.6;n=3940 | 5.6(5.1-6.6);31.1;n=20409 | <0.0001*** | 6.8(5.6-9.3);70.6;n=10506 | 5.4(4.95-6.1);20.2;n=13843 | <0.0001*** |
| TyG | 8.41(8.2-8.73);11.61;n=3940 | 7.17(6.8-7.55);8.05;n=20409 | <0.0001*** | 7.9(7.65-8.26);11.61;n=10506 | 6.94(6.65-7.19);7.43;n=13843 | <0.0001*** |

DM: diabetes mellitus; HF: heart failure; AF: atrial fibrillation; AMI: acute myocardial infarction; VTF: ventricular tachycardia/fibrillation; IHD: ischemic heart disease; COPD: chronic obstructive pulmonary disease; PVD: peripheral vascular disease; TIA: transient ischemic attack; ACEI: angiotensinogen converting enzyme inhibitor; ARB: angiotensin receptor blocker; TyG: triglyceride-glucose

**Supplementary Table 5. Basic characteristics of TyG index cut-off with new onset DM and new onset HF**

* for p≤ 0.05, ** for p ≤ 0.01, *** for p ≤ 0.001

| **Characteristics** | **TyG>7.68 (N=7421) Median (IQR);Max;N or Count(%)** | **TyG**$\boldsymbol{\leq}$**7.68 (N=16928) Median (IQR);Max;N or Count(%)** | **P value** | **TyG>7.24 (N=13147) Median (IQR);Max;N or Count(%)** | **TyG**$\boldsymbol{\leq}$**7.24 (N=11202) Median (IQR);Max;N or Count(%)** | **P value** |
| --- | --- | --- | --- | --- | --- | --- |
| Male gender | 3140(42.31%) | 6649(39.27%) | 0.0040** | 5473(41.62%) | 4316(38.52%) | 0.0014** |
| Baseline age, year | 63.58(52.86-71.54);98.94;n=7421 | 61.94(50.66-71.31);100.97;n=16928 | <0.0001*** | 63.34(52.48-71.46);98.94;n=13147 | 61.3(50.06-71.29);100.97;n=11202 | <0.0001*** |
| Charlson score | 2.0(1.0-3.0);9.0;n=7421 | 2.0(1.0-3.0);11.0;n=16928 | <0.0001*** | 2.0(1.0-3.0);9.0;n=13147 | 2.0(1.0-3.0);11.0;n=11202 | <0.0001*** |
| Liver diseases | 2(0.02%) | 5(0.02%) | 0.7634 | 4(0.03%) | 3(0.02%) | 0.832 |
| Endocrine | 9(0.12%) | 22(0.12%) | 0.9836 | 15(0.11%) | 16(0.14%) | 0.6558 |
| Hypertension | 305(4.10%) | 737(4.35%) | 0.4267 | 569(4.32%) | 473(4.22%) | 0.7213 |
| IHD | 73(0.98%) | 168(0.99%) | 0.994 | 127(0.96%) | 114(1.01%) | 0.7361 |
| COPD | 5(0.06%) | 26(0.15%) | 0.1237 | 10(0.07%) | 21(0.18%) | 0.0247* |
| Gastrointestinal | 166(2.23%) | 420(2.48%) | 0.2835 | 294(2.23%) | 292(2.60%) | 0.073 |
| PVD | 3(0.04%) | 10(0.05%) | 0.7809 | 5(0.03%) | 8(0.07%) | 0.3981 |
| TIA/Ischemic stroke | 28(0.37%) | 70(0.41%) | 0.7648 | 59(0.44%) | 39(0.34%) | 0.2587 |
| Gastrointestinal bleeding | 35(0.47%) | 79(0.46%) | 0.96 | 63(0.47%) | 51(0.45%) | 0.8594 |
| Cancer | 50(0.67%) | 111(0.65%) | 0.9418 | 84(0.63%) | 77(0.68%) | 0.7021 |
| ACEI | 724(9.75%) | 1100(6.49%) | <0.0001*** | 1199(9.11%) | 625(5.57%) | <0.0001*** |
| ARB | 35(0.47%) | 54(0.31%) | 0.0902 | 54(0.41%) | 35(0.31%) | 0.2478 |
| Calcium channel blockers | 964(12.99%) | 1804(10.65%) | <0.0001*** | 1657(12.60%) | 1111(9.91%) | <0.0001*** |
| Beta blockers | 920(12.39%) | 1700(10.04%) | <0.0001*** | 1596(12.13%) | 1024(9.14%) | <0.0001*** |
| Nitrates | 405(5.45%) | 716(4.22%) | <0.0001*** | 688(5.23%) | 433(3.86%) | <0.0001*** |
| Antihypertensive drugs | 325(4.37%) | 731(4.31%) | 0.8631 | 584(4.44%) | 472(4.21%) | 0.4213 |
| Anti-Diabetic drugs | 677(9.12%) | 652(3.85%) | <0.0001*** | 978(7.43%) | 351(3.13%) | <0.0001*** |
| Statins and fibrates | 739(9.95%) | 1133(6.69%) | <0.0001*** | 1185(9.01%) | 687(6.13%) | <0.0001*** |
| Diuretics for hypertension | 468(6.30%) | 875(5.16%) | 0.0008*** | 820(6.23%) | 523(4.66%) | <0.0001*** |
| Diuretics for heart failure | 128(1.72%) | 229(1.35%) | 0.0330* | 212(1.61%) | 145(1.29%) | 0.0482* |
| Anticoagulants | 28(0.37%) | 27(0.15%) | 0.0017** | 42(0.31%) | 13(0.11%) | 0.0014** |
| Antiplatelets | 490(6.60%) | 866(5.11%) | <0.0001*** | 833(6.33%) | 523(4.66%) | <0.0001*** |
| Lipid-lowering drugs | 554(7.46%) | 943(5.57%) | <0.0001*** | 939(7.14%) | 558(4.98%) | <0.0001*** |
| Mean corpuscular volume, fL | 89.1(85.7-92.1);108.0;n=3074 | 89.9(86.7-92.8);132.3;n=6792 | <0.0001*** | 89.3(85.9-92.2);132.3;n=5367 | 90.2(86.9-93.0);119.5;n=4499 | <0.0001*** |
| Lymphocyte, x10^9/L | 1.92(1.5-2.5);5.5;n=1778 | 1.8(1.4-2.2);85.28;n=3824 | <0.0001*** | 1.9(1.5-2.4);5.5;n=3048 | 1.8(1.3-2.2);85.28;n=2554 | <0.0001*** |
| Metamyelocyte, x10^9/L | 0.19(0.12-0.62);2.0;n=25 | 0.18(0.06-0.58);1.47;n=20 | 0.5371 | 0.2(0.12-0.63);2.0;n=33 | 0.1(0.04-0.4);1.1;n=12 | 0.1029 |
| Monocyte, x10^9/L | 0.5(0.4-0.69);3.22;n=1774 | 0.5(0.4-0.6);3.18;n=3818 | <0.0001*** | 0.5(0.4-0.6);3.22;n=3043 | 0.5(0.33-0.6);3.18;n=2549 | <0.0001*** |
| Neutrophil, x10^9/L | 4.6(3.6-6.3);28.2;n=1774 | 4.1(3.2-5.7);39.48;n=3816 | <0.0001*** | 4.5(3.5-6.2);39.48;n=3043 | 4.1(3.1-5.6);36.67;n=2547 | <0.0001*** |
| White blood count, x10^9/L | 7.61(6.32-9.4);30.8;n=3094 | 6.9(5.7-8.4);91.7;n=6830 | <0.0001*** | 7.5(6.2-9.2);47.0;n=5406 | 6.7(5.57-8.1);91.7;n=4518 | <0.0001*** |
| Mean cell haemoglobin, pg | 30.5(29.2-31.7);38.0;n=3074 | 30.6(29.3-31.7);44.1;n=6792 | 0.0442* | 30.5(29.2-31.6);44.1;n=5367 | 30.7(29.4-31.7);41.5;n=4499 | <0.0001*** |
| Myelocyte, x10^9/L | 0.32(0.14-0.8);2.87;n=14 | 0.13(0.1-0.4);0.94;n=16 | 0.2794 | 0.32(0.12-0.8);2.87;n=20 | 0.12(0.1-0.34);0.81;n=10 | 0.2523 |
| Platelet, x10^9/L | 240.0(203.0-285.0);952.0;n=3094 | 235.0(196.0-280.0);1250.0;n=6830 | 0.0001*** | 240.0(201.0-284.5);1250.0;n=5406 | 233.0(195.0-277.0);957.0;n=4518 | <0.0001*** |
| Reticulocyte, x10^9/L | 71.04(46.2-92.7);307.58;n=121 | 52.44(37.11-80.6);324.0;n=247 | 0.0002*** | 67.59(43.94-90.85);307.58;n=202 | 50.98(34.32-78.04);324.0;n=166 | 0.0006*** |
| Red blood count, x10^12/L | 4.49(4.15-4.89);7.62;n=3074 | 4.41(4.09-4.75);7.61;n=6789 | <0.0001*** | 4.49(4.14-4.86);7.62;n=5367 | 4.37(4.07-4.71);7.61;n=4496 | <0.0001*** |
| Hematocrit, L/L | 0.4(0.37-0.43);0.551;n=432 | 0.39(0.36-0.42);0.519;n=936 | 0.0641 | 0.4(0.37-0.42);0.551;n=734 | 0.39(0.36-0.42);0.519;n=634 | 0.0116* |
| K/Potassium, mmol/L | 4.2(3.87-4.51);6.7;n=7294 | 4.2(3.9-4.5);13.3;n=16318 | 0.8109 | 4.2(3.88-4.5);10.0;n=12856 | 4.2(3.9-4.5);13.3;n=10756 | 0.0578 |
| Albumin, g/L | 42.0(39.9-44.0);56.0;n=4108 | 42.0(39.9-44.0);58.0;n=9259 | 0.0113* | 42.0(40.0-44.0);56.0;n=7319 | 42.0(39.6-44.0);58.0;n=6048 | <0.0001*** |
| Na/Sodium, mmol/L | 140.0(138.0-142.0);168.0;n=7307 | 141.0(139.0-142.0);158.0;n=16363 | <0.0001*** | 140.0(138.3-142.0);168.0;n=12883 | 141.0(139.0-142.0);152.0;n=10787 | <0.0001*** |
| Urea, mmol/L | 5.6(4.7-6.8);46.4;n=7308 | 5.5(4.6-6.6);47.3;n=16357 | <0.0001*** | 5.6(4.6-6.8);46.4;n=12883 | 5.5(4.6-6.6);47.3;n=10782 | <0.0001*** |
| Protein, g/L | 75.0(72.0-79.0);109.0;n=4093 | 74.4(71.0-78.0);109.0;n=9212 | <0.0001*** | 75.0(72.0-78.5);109.0;n=7290 | 74.0(71.0-77.3);100.0;n=6015 | <0.0001*** |
| Creatinine, umol/L | 85.0(73.0-100.0);1509.0;n=7319 | 83.0(71.0-97.0);807.0;n=16392 | <0.0001*** | 85.0(73.0-99.0);1509.0;n=12905 | 82.0(71.0-96.0);629.0;n=10806 | <0.0001*** |
| Alkaline phosphatase, U/L | 81.0(67.0-98.0);885.0;n=3836 | 76.0(62.0-93.0);827.0;n=8631 | <0.0001*** | 80.0(66.0-97.0);885.0;n=6835 | 74.5(61.0-91.0);624.0;n=5632 | <0.0001*** |
| Aspartate transaminase, U/L | 23.0(18.0-31.0);2640.0;n=781 | 22.0(18.0-28.0);1787.0;n=1659 | 0.0061** | 23.0(18.0-30.0);2640.0;n=1338 | 22.0(18.0-28.0);1516.0;n=1102 | 0.0655 |
| Alanine transaminase, U/L | 23.0(17.0-34.0);918.0;n=3143 | 20.0(14.0-28.0);700.0;n=7273 | <0.0001*** | 22.83(16.0-33.0);918.0;n=5656 | 19.0(14.0-27.0);700.0;n=4760 | <0.0001*** |
| Bilirubin, umol/L | 9.2(7.0-12.4);107.0;n=3869 | 10.0(7.4-13.0);216.0;n=8698 | <0.0001*** | 9.5(7.0-12.7);140.0;n=6891 | 10.0(7.4-13.0);216.0;n=5676 | 0.0003*** |
| Triglyceride, mmol/L | 2.44(1.76-3.24);38.3;n=7421 | 1.18(0.88-1.55);3.36;n=16928 | <0.0001*** | 1.96(1.52-2.63);38.3;n=13147 | 1.01(0.78-1.25);3.0;n=11202 | <0.0001*** |
| Low-density lipoprotein, mmol/L | 3.13(2.5-3.81);8.0052;n=6280 | 3.18(2.57-3.82);9.3127;n=14459 | 0.0026** | 3.21(2.57-3.88);9.3045;n=11249 | 3.12(2.54-3.76);9.3127;n=9490 | <0.0001*** |
| High-density lipoprotein, mmol/L | 1.14(0.98-1.35);3.38;n=6665 | 1.37(1.16-1.64);4.14;n=14519 | <0.0001*** | 1.19(1.01-1.4);3.38;n=11655 | 1.44(1.21-1.72);4.14;n=9529 | <0.0001*** |
| Cholesterol, mmol/L | 5.54(4.86-6.29);19.04;n=7406 | 5.2(4.57-5.92);12.205;n=16899 | <0.0001*** | 5.5(4.8-6.21);19.04;n=13121 | 5.11(4.5-5.8);11.5;n=11184 | <0.0001*** |
| HbA1c, g/dL | 13.5(12.5-14.5);19.5;n=2900 | 13.3(12.3-14.1);20.0;n=6385 | <0.0001*** | 13.4(12.4-14.4);20.0;n=5047 | 13.2(12.3-14.1);19.5;n=4238 | <0.0001*** |
| Fast glucose, mmol/L | 7.23(5.8-10.12);70.6;n=7421 | 5.5(5.0-6.3);23.1;n=16928 | <0.0001*** | 6.5(5.5-8.7);70.6;n=13147 | 5.3(4.9-5.94);20.2;n=11202 | <0.0001*** |
| TyG | 8.08(7.86-8.44);11.61;n=7421 | 7.05(6.72-7.35);7.68;n=16928 | <0.0001*** | 7.76(7.48-8.15);11.61;n=13147 | 6.84(6.56-7.05);7.24;n=11202 | <0.0001*** |

DM: diabetes mellitus; HF: heart failure; AF: atrial fibrillation; AMI: acute myocardial infarction; VTF: ventricular tachycardia/fibrillation; IHD: ischemic heart disease; COPD: chronic obstructive pulmonary disease; PVD: peripheral vascular disease; TIA: transient ischemic attack; ACEI: angiotensinogen converting enzyme inhibitor; ARB: angiotensin receptor blocker; TyG: triglyceride-glucose

**Supplementary Table 6. Basic characteristics of TyG index cut-off with new onset AF, new onset AMI and new onset VTF**

* for p≤ 0.05, ** for p ≤ 0.01, *** for p ≤ 0.001

| **Characteristics** | **TyG>7.75 (N=6667) Median (IQR);Max;N or Count(%)** | **TyG**$\boldsymbol{\leq}$**7.75 (N=17682) Median (IQR);Max;N or Count(%)** | **P value** | **TyG>7.66 (N=7651) Median (IQR);Max;N or Count(%)** | **TyG**$\boldsymbol{\leq}$**7.66 (N=16698) Median (IQR);Max;N or Count(%)** | **P value** | **TyG>7.79 (N=6244) Median (IQR);Max;N or Count(%)** | **TyG**$\boldsymbol{\leq}$**7.79 (N=18105) Median (IQR);Max;N or Count(%)** | **P value** |
| --- | --- | --- | --- | --- | --- | --- | --- | --- | --- |
| Male gender | 2843(42.64%) | 6946(39.28%) | 0.0020** | 3229(42.20%) | 6560(39.28%) | 0.0052** | 2669(42.74%) | 7120(39.32%) | 0.0021** |
| Baseline age, year | 63.66(52.87-71.57);98.94;n=6667 | 61.98(50.73-71.31);100.97;n=17682 | <0.0001*** | 63.52(52.79-71.53);98.94;n=7651 | 61.94(50.66-71.31);100.97;n=16698 | <0.0001*** | 63.66(52.88-71.5);98.94;n=6244 | 62.03(50.77-71.33);100.97;n=18105 | <0.0001*** |
| Charlson score | 2.0(1.0-3.0);9.0;n=6667 | 2.0(1.0-3.0);11.0;n=17682 | <0.0001*** | 2.0(1.0-3.0);9.0;n=7651 | 2.0(1.0-3.0);11.0;n=16698 | <0.0001*** | 2.0(1.0-3.0);9.0;n=6244 | 2.0(1.0-3.0);11.0;n=18105 | <0.0001*** |
| Liver diseases | 2(0.02%) | 5(0.02%) | 0.7239 | 2(0.02%) | 5(0.02%) | 0.8067 | 2(0.03%) | 5(0.02%) | 0.7984 |
| Endocrine | 9(0.13%) | 22(0.12%) | 0.9964 | 9(0.11%) | 22(0.13%) | 0.926 | 8(0.12%) | 23(0.12%) | 0.8532 |
| Hypertension | 271(4.06%) | 771(4.36%) | 0.3479 | 320(4.18%) | 722(4.32%) | 0.652 | 253(4.05%) | 789(4.35%) | 0.3413 |
| IHD | 61(0.91%) | 180(1.01%) | 0.5192 | 77(1.00%) | 164(0.98%) | 0.9156 | 60(0.96%) | 181(0.99%) | 0.849 |
| COPD | 5(0.07%) | 26(0.14%) | 0.2291 | 6(0.07%) | 25(0.14%) | 0.2102 | 5(0.08%) | 26(0.14%) | 0.314 |
| Gastrointestinal | 147(2.20%) | 439(2.48%) | 0.2359 | 171(2.23%) | 415(2.48%) | 0.2668 | 138(2.21%) | 448(2.47%) | 0.2713 |
| PVD | 3(0.04%) | 10(0.05%) | 0.9706 | 3(0.03%) | 10(0.05%) | 0.7269 | 3(0.04%) | 10(0.05%) | 0.9158 |
| TIA/Ischemic stroke | 26(0.38%) | 72(0.40%) | 0.9403 | 30(0.39%) | 68(0.40%) | 0.9495 | 23(0.36%) | 75(0.41%) | 0.7068 |
| Gastrointestinal bleeding | 27(0.40%) | 87(0.49%) | 0.4366 | 36(0.47%) | 78(0.46%) | 0.9481 | 25(0.40%) | 89(0.49%) | 0.4245 |
| Cancer | 48(0.71%) | 113(0.63%) | 0.5477 | 52(0.67%) | 109(0.65%) | 0.878 | 44(0.70%) | 117(0.64%) | 0.691 |
| ACEI | 657(9.85%) | 1167(6.59%) | <0.0001*** | 738(9.64%) | 1086(6.50%) | <0.0001*** | 628(10.05%) | 1196(6.60%) | <0.0001*** |
| ARB | 31(0.46%) | 58(0.32%) | 0.146 | 35(0.45%) | 54(0.32%) | 0.1366 | 31(0.49%) | 58(0.32%) | 0.063 |
| Calcium channel blockers | 865(12.97%) | 1903(10.76%) | <0.0001*** | 994(12.99%) | 1774(10.62%) | <0.0001*** | 813(13.02%) | 1955(10.79%) | <0.0001*** |
| Beta blockers | 832(12.47%) | 1788(10.11%) | <0.0001*** | 942(12.31%) | 1678(10.04%) | <0.0001*** | 782(12.52%) | 1838(10.15%) | <0.0001*** |
| Nitrates | 369(5.53%) | 752(4.25%) | <0.0001*** | 414(5.41%) | 707(4.23%) | 0.0001*** | 354(5.66%) | 767(4.23%) | <0.0001*** |
| Antihypertensive drugs | 294(4.40%) | 762(4.30%) | 0.7697 | 334(4.36%) | 722(4.32%) | 0.9142 | 276(4.42%) | 780(4.30%) | 0.7468 |
| Anti-Diabetic drugs | 625(9.37%) | 704(3.98%) | <0.0001*** | 688(8.99%) | 641(3.83%) | <0.0001*** | 602(9.64%) | 727(4.01%) | <0.0001*** |
| Statins and fibrates | 682(10.22%) | 1190(6.73%) | <0.0001*** | 761(9.94%) | 1111(6.65%) | <0.0001*** | 642(10.28%) | 1230(6.79%) | <0.0001*** |
| Diuretics for hypertension | 427(6.40%) | 916(5.18%) | 0.0005*** | 476(6.22%) | 867(5.19%) | 0.0023** | 401(6.42%) | 942(5.20%) | 0.0007*** |
| Diuretics for heart failure | 118(1.76%) | 239(1.35%) | 0.0201* | 132(1.72%) | 225(1.34%) | 0.0289* | 110(1.76%) | 247(1.36%) | 0.0310* |
| Anticoagulants | 26(0.38%) | 29(0.16%) | 0.0016** | 28(0.36%) | 27(0.16%) | 0.0030** | 25(0.40%) | 30(0.16%) | 0.0014** |
| Antiplatelets | 446(6.68%) | 910(5.14%) | <0.0001*** | 505(6.60%) | 851(5.09%) | <0.0001*** | 420(6.72%) | 936(5.16%) | <0.0001*** |
| Lipid-lowering drugs | 502(7.52%) | 995(5.62%) | <0.0001*** | 571(7.46%) | 926(5.54%) | <0.0001*** | 474(7.59%) | 1023(5.65%) | <0.0001*** |
| Mean corpuscular volume, fL | 89.2(85.75-92.1);108.0;n=2763 | 89.9(86.6-92.8);132.3;n=7103 | <0.0001*** | 89.1(85.7-92.1);108.0;n=3163 | 89.9(86.7-92.8);132.3;n=6703 | <0.0001*** | 89.2(85.7-92.2);108.0;n=2579 | 89.9(86.6-92.8);132.3;n=7287 | <0.0001*** |
| Lymphocyte, x10^9/L | 2.0(1.5-2.5);5.2;n=1599 | 1.8(1.4-2.3);85.28;n=4003 | <0.0001*** | 1.94(1.5-2.5);5.5;n=1817 | 1.8(1.4-2.2);85.28;n=3785 | <0.0001*** | 2.0(1.5-2.5);5.2;n=1495 | 1.8(1.4-2.3);85.28;n=4107 | <0.0001*** |
| Metamyelocyte, x10^9/L | 0.19(0.12-0.56);2.0;n=21 | 0.18(0.08-0.64);1.79;n=24 | 0.8644 | 0.2(0.12-0.62);2.0;n=26 | 0.18(0.06-0.52);1.47;n=19 | 0.3116 | 0.18(0.1-0.59);2.0;n=20 | 0.19(0.09-0.63);1.79;n=25 | 0.9362 |
| Monocyte, x10^9/L | 0.5(0.4-0.69);3.22;n=1595 | 0.5(0.4-0.6);3.18;n=3997 | <0.0001*** | 0.5(0.4-0.69);3.22;n=1813 | 0.5(0.4-0.6);3.18;n=3779 | <0.0001*** | 0.5(0.4-0.7);3.22;n=1491 | 0.5(0.4-0.6);3.18;n=4101 | <0.0001*** |
| Neutrophil, x10^9/L | 4.6(3.6-6.3);28.2;n=1595 | 4.2(3.2-5.7);39.48;n=3995 | <0.0001*** | 4.6(3.6-6.4);28.2;n=1813 | 4.1(3.2-5.7);39.48;n=3777 | <0.0001*** | 4.6(3.6-6.4);28.2;n=1491 | 4.2(3.2-5.7);39.48;n=4099 | <0.0001*** |
| White blood count, x10^9/L | 7.63(6.33-9.4);30.8;n=2780 | 6.9(5.7-8.44);91.7;n=7144 | <0.0001*** | 7.63(6.3-9.4);30.8;n=3185 | 6.86(5.7-8.4);91.7;n=6739 | <0.0001*** | 7.66(6.34-9.4);30.8;n=2595 | 6.9(5.7-8.5);91.7;n=7329 | <0.0001*** |
| Mean cell haemoglobin, pg | 30.5(29.2-31.7);38.0;n=2763 | 30.6(29.3-31.7);44.1;n=7103 | 0.0974 | 30.5(29.2-31.7);38.0;n=3163 | 30.6(29.4-31.7);44.1;n=6703 | 0.0156* | 30.5(29.2-31.7);38.0;n=2579 | 30.6(29.3-31.7);44.1;n=7287 | 0.1618 |
| Myelocyte, x10^9/L | 0.3(0.13-0.78);0.97;n=13 | 0.14(0.1-0.45);2.87;n=17 | 0.63 | 0.34(0.14-0.8);2.87;n=15 | 0.12(0.1-0.4);0.94;n=15 | 0.2207 | 0.32(0.09-0.8);0.97;n=12 | 0.15(0.11-0.4);2.87;n=18 | 0.5818 |
| Platelet, x10^9/L | 240.0(203.0-285.0);952.0;n=2780 | 235.0(196.0-280.0);1250.0;n=7144 | 0.0005*** | 241.0(203.0-285.0);952.0;n=3185 | 235.0(196.0-280.0);1250.0;n=6739 | <0.0001*** | 240.0(203.0-285.0);952.0;n=2595 | 236.0(196.0-280.0);1250.0;n=7329 | 0.002** |
| Reticulocyte, x10^9/L | 71.53(47.5-93.64);307.58;n=112 | 52.47(36.95-80.76);324.0;n=256 | 0.0001*** | 71.0(46.75-92.38);307.58;n=122 | 52.37(37.11-80.6);324.0;n=246 | 0.0002*** | 73.27(48.35-95.16);307.58;n=102 | 52.37(37.22-80.6);324.0;n=266 | <0.0001*** |
| Red blood count, x10^12/L | 4.5(4.16-4.9);7.62;n=2763 | 4.41(4.09-4.75);7.61;n=7100 | <0.0001*** | 4.49(4.15-4.89);7.62;n=3163 | 4.4(4.09-4.75);7.61;n=6700 | <0.0001*** | 4.5(4.15-4.9);7.62;n=2579 | 4.41(4.09-4.76);7.61;n=7284 | <0.0001*** |
| Hematocrit, L/L | 0.4(0.37-0.43);0.551;n=379 | 0.39(0.36-0.42);0.519;n=989 | 0.0115* | 0.4(0.37-0.43);0.551;n=445 | 0.39(0.36-0.42);0.519;n=923 | 0.0302* | 0.4(0.37-0.43);0.551;n=351 | 0.39(0.36-0.42);0.519;n=1017 | 0.0037** |
| K/Potassium, mmol/L | 4.2(3.86-4.5);6.7;n=6553 | 4.2(3.9-4.5);13.3;n=17059 | 0.6731 | 4.2(3.87-4.51);6.7;n=7519 | 4.2(3.9-4.5);13.3;n=16093 | 0.8575 | 4.2(3.85-4.5);6.7;n=6134 | 4.2(3.9-4.5);13.3;n=17478 | 0.4789 |
| Albumin, g/L | 42.0(40.0-44.0);56.0;n=3691 | 42.0(39.8-44.0);58.0;n=9676 | 0.0267* | 42.0(39.9-44.0);56.0;n=4237 | 42.0(39.8-44.0);58.0;n=9130 | 0.0044** | 42.0(40.0-44.0);56.0;n=3449 | 42.0(39.8-44.0);58.0;n=9918 | 0.0696 |
| Na/Sodium, mmol/L | 140.0(138.0-142.0);168.0;n=6563 | 141.0(139.0-142.0);158.0;n=17107 | <0.0001*** | 140.0(138.0-142.0);168.0;n=7532 | 141.0(139.0-142.0);158.0;n=16138 | <0.0001*** | 140.0(138.0-142.0);168.0;n=6144 | 141.0(139.0-142.0);158.0;n=17526 | <0.0001*** |
| Urea, mmol/L | 5.6(4.7-6.88);46.4;n=6564 | 5.5(4.6-6.6);47.3;n=17101 | <0.0001*** | 5.6(4.7-6.8);46.4;n=7534 | 5.5(4.6-6.6);47.3;n=16131 | <0.0001*** | 5.6(4.7-6.9);46.4;n=6145 | 5.5(4.6-6.6);47.3;n=17520 | <0.0001*** |
| Protein, g/L | 75.0(72.0-79.0);109.0;n=3676 | 74.4(71.0-78.0);109.0;n=9629 | <0.0001*** | 75.0(72.0-79.0);109.0;n=4222 | 74.3(71.0-78.0);109.0;n=9083 | <0.0001*** | 75.1(72.0-79.0);109.0;n=3436 | 74.5(71.0-78.0);109.0;n=9869 | <0.0001*** |
| Creatinine, umol/L | 86.0(73.0-100.0);1509.0;n=6575 | 83.0(71.0-97.0);1063.0;n=17136 | <0.0001*** | 85.0(73.0-100.0);1509.0;n=7545 | 83.0(71.0-97.0);807.0;n=16166 | <0.0001*** | 86.0(74.0-100.0);1509.0;n=6156 | 83.0(71.0-97.0);1476.0;n=17555 | <0.0001*** |
| Alkaline phosphatase, U/L | 81.0(67.0-98.0);885.0;n=3449 | 76.0(62.0-93.0);827.0;n=9018 | <0.0001*** | 81.0(67.0-98.0);885.0;n=3959 | 76.0(62.0-93.0);827.0;n=8508 | <0.0001*** | 81.0(67.0-99.0);885.0;n=3221 | 76.0(62.0-93.0);827.0;n=9246 | <0.0001*** |
| Aspartate transaminase, U/L | 23.0(18.0-31.0);2640.0;n=703 | 22.0(18.0-28.0);1787.0;n=1737 | 0.0093** | 23.0(18.0-31.0);2640.0;n=805 | 22.0(18.0-28.0);1787.0;n=1635 | 0.0054** | 23.0(18.5-31.0);2640.0;n=650 | 22.0(18.0-28.0);1787.0;n=1790 | 0.0178* |
| Alanine transaminase, U/L | 23.0(17.0-34.0);918.0;n=2805 | 20.0(14.0-29.0);700.0;n=7611 | <0.0001*** | 23.0(17.0-34.0);918.0;n=3243 | 20.0(14.0-28.0);700.0;n=7173 | <0.0001*** | 23.0(17.0-34.0);918.0;n=2630 | 20.0(14.0-29.0);700.0;n=7786 | <0.0001*** |
| Bilirubin, umol/L | 9.3(7.0-12.35);107.0;n=3475 | 10.0(7.3-13.0);216.0;n=9092 | <0.0001*** | 9.2(7.0-12.5);107.0;n=3992 | 10.0(7.4-13.0);216.0;n=8575 | <0.0001*** | 9.2(7.0-12.2);107.0;n=3245 | 10.0(7.3-13.0);216.0;n=9322 | <0.0001*** |
| Triglyceride, mmol/L | 2.51(1.79-3.36);38.3;n=6667 | 1.2(0.89-1.59);3.85;n=17682 | <0.0001*** | 2.42(1.76-3.2);38.3;n=7651 | 1.17(0.87-1.53);3.36;n=16698 | <0.0001*** | 2.57(1.81-3.44);38.3;n=6244 | 1.21(0.9-1.62);6.2;n=18105 | <0.0001*** |
| Low-density lipoprotein, mmol/L | 3.13(2.49-3.81);8.0052;n=5619 | 3.18(2.57-3.82);9.3127;n=15120 | 0.0002*** | 3.15(2.51-3.82);8.0052;n=6467 | 3.18(2.57-3.82);9.3127;n=14272 | 0.0242* | 3.11(2.48-3.79);8.0052;n=5241 | 3.19(2.58-3.82);9.3127;n=15498 | <0.0001*** |
| High-density lipoprotein, mmol/L | 1.14(0.98-1.34);3.38;n=6001 | 1.37(1.15-1.63);4.14;n=15183 | <0.0001*** | 1.15(0.98-1.35);3.38;n=6855 | 1.38(1.16-1.65);4.14;n=14329 | <0.0001*** | 1.13(0.97-1.34);3.38;n=5619 | 1.36(1.14-1.63);4.14;n=15565 | <0.0001*** |
| Cholesterol, mmol/L | 5.55(4.88-6.3);19.04;n=6652 | 5.22(4.59-5.94);12.205;n=17653 | <0.0001*** | 5.54(4.86-6.29);19.04;n=7635 | 5.2(4.57-5.91);12.205;n=16670 | <0.0001*** | 5.56(4.88-6.3);19.04;n=6231 | 5.23(4.59-5.94);12.205;n=18074 | <0.0001*** |
| HbA1c, g/dL | 13.5(12.5-14.5);19.5;n=2607 | 13.3(12.3-14.1);20.0;n=6678 | <0.0001*** | 13.5(12.5-14.5);19.5;n=2983 | 13.3(12.3-14.1);20.0;n=6302 | <0.0001*** | 13.5(12.5-14.5);19.5;n=2434 | 13.3(12.3-14.2);20.0;n=6851 | <0.0001*** |
| Fast glucose, mmol/L | 7.4(5.8-10.4);70.6;n=6667 | 5.5(5.0-6.4);30.2;n=17682 | <0.0001*** | 7.2(5.7-10.1);70.6;n=7651 | 5.5(5.0-6.3);23.1;n=16698 | <0.0001*** | 7.6(5.81-10.66);70.6;n=6244 | 5.5(5.0-6.4);31.1;n=18105 | <0.0001*** |
| TyG | 8.14(7.92-8.49);11.61;n=6667 | 7.08(6.74-7.39);7.75;n=17682 | <0.0001*** | 8.07(7.85-8.42);11.61;n=7651 | 7.04(6.72-7.34);7.66;n=16698 | <0.0001*** | 8.17(7.96-8.52);11.61;n=6244 | 7.09(6.75-7.42);7.79;n=18105 | <0.0001*** |

DM: diabetes mellitus; HF: heart failure; AF: atrial fibrillation; AMI: acute myocardial infarction; VTF: ventricular tachycardia/fibrillation; IHD: ischemic heart disease; COPD: chronic obstructive pulmonary disease; PVD: peripheral vascular disease; TIA: transient ischemic attack; ACEI: angiotensinogen converting enzyme inhibitor; ARB: angiotensin receptor blocker; TyG: triglyceride-glucose
